# Trans-ancestry genome-wide association meta-analysis of antidepressant response to selective serotonin reuptake inhibitors in clinical studies of depression

**DOI:** 10.64898/2026.06.03.26354703

**Authors:** Ke Hu, Chris Wai Hang Lo, Swapnil Awasthi, Oliver Pain, Madhurbain Singh, Yeeun Ahn, Katherine J Aitchison, Bernhard T Baune, Joanna M Biernacka, Guido Bondolfi, Tania Carrillo-Roa, Hong Choi, Darina Czamara, Katharina Domschke, Chiara Fabbri, Steven P Hamilton, Marcus Ising, Yoonjeong Jang, Masaki Kato, Doh Kwan Kim, Dongjun Kim, Byung-Chul Lee, Glyn Lewis, Shinn-Won Lim, Yu-Li Liu, Woojae Myung, Nader Perroud, Alessandro Serretti, Shih-Jen Tsai, Rudolf Uher, Richard Weinshilboum, Hong-Hee Won, Major Depressive Disorder Working Group of the Psychiatric Genomics Consortium, Stephan Ripke, Jonathan RI Coleman, Cathryn M Lewis

## Abstract

Antidepressants are widely prescribed for major depressive disorder, yet only one-third of patients achieve remission after initial treatment. Previous genome-wide association studies (GWAS) of clinically assessed antidepressant response combined multiple antidepressant classes, potentially obscuring class-specific effects. This study focused on selective serotonin reuptake inhibitors (SSRIs), often first-line due to better tolerability. Data from 15 cohorts across four ancestries were integrated: European (N = 3887; 11 studies), East Asian (N = 1068; 4), African (N = 277; 1), and Admixed American (N = 250; 1). GWAS of non-remission and percentage improvement were conducted within cohorts, followed by ancestry-specific meta-analyses and trans-ancestry meta-regression. Single nucleotide polymorphism (SNP)-based heritability was estimated in European samples. Polygenic scores were used for leave-one-out prediction and to assess shared genetic architecture with psychiatric traits. Gene-level and gene-set enrichment analyses were also performed. No genome-wide significant variants were identified for either outcome in any ancestry-specific or trans-ancestry analyses. However, trans-ancestry meta-regression yielded eight independent loci with suggestive associations (p < 1 × 10^−5^) for non-remission and 17 for percentage improvement. Gene-set analyses revealed nominal enrichment of the serotonergic synapse pathway for non-remission. SNP-based heritability estimates were not significantly different from zero for either outcome. Better SSRI response was nominally associated with lower genetic predisposition to major depressive disorder, post-traumatic stress disorder, and schizophrenia. This study represents the largest trans-ancestry GWAS of SSRI response, highlighting emerging biological signals. Limited power emphasises the need for larger and ancestrally diverse cohorts to better characterise the genetic architecture of antidepressant response.

## 1 Introduction

Major Depressive Disorder (MDD) is a leading cause of global health-related burden and disability, with an estimated lifetime prevalence of 15% [1]. In the UK, psychological and psychosocial therapy are considered as first-line treatment for patients with subthreshold or mild depression, with antidepressants recommended for those with moderate to severe depression [2]. Selective serotonin reuptake inhibitors (SSRIs) are the most commonly prescribed pharmacological treatment for MDD, largely due to their selectivity for the serotonergic pathway [3], making them more tolerable than other antidepressant classes, such as serotonin noradrenaline reuptake inhibitors (SNRIs) or tricyclic antidepressants (TCAs) [4,5]. However, it remains unclear whether SSRIs are more efficacious than these alternative classes [6].

Although antidepressants show better efficacy compared with placebo in reducing depressive symptoms, only one-third of patients with MDD achieve remission after their initial treatment [6,7]. Therefore, it is of critical importance to investigate the factors underlying individual variability in antidepressant response and advance personalised treatment strategies.

One source of individual differences in antidepressant response is common genetic variation. Genetic polymorphisms in genes encoding drug metabolising enzymes, particularly *CYP2C19* and *CYP2D6*, are used in pharmacogenetic guidelines for antidepressant prescribing and dosing, but only explain part of variation in treatment response [8,9]. Previous GWAS of antidepressant response by the Psychiatric Genomics Consortium (PGC) MDD Working Group, including 13 clinical studies and 5843 MDD patients, revealed no genome-wide significant SNPs but estimated a SNP-based heritability of 13%, suggesting a moderate contribution of common genetic variation [10].

The genetic basis of SSRI response may have a distinct profile compared to the broader antidepressant response, which encompasses heterogeneity across classes of antidepressants. However, most GWAS of clinically assessed SSRI response have lacked sufficient statistical power and have failed to identify significant common genetic variants associated with response [11–13]. Fabbri et al. [14] increased variant coverage and identified two significant loci in a meta-analysis of Genome-Based Therapeutic Drugs for Depression (GENDEP) and Sequenced Treatment Alternatives to Relieve Depression (STAR*D) studies, but findings were only partially replicated in the Pharmacogenomics Research Network Antidepressant Medication Pharmacogenomics Study (PGRN-AMPS) and the NEWMEDS consortium. These studies highlight the need for further investigation with larger and more diverse cohorts, as no large-scale GWAS of SSRI response in clinical studies has yet been performed.

Alternatively, broader SSRI response phenotypes derived from retrospective self-reported questionnaires or inferred from drug switching patterns in prescription registries have been analysed to increase the sample sizes and improve statistical power, but with limited findings. The 23andMe Research Team (N = 19,740) identified one significant locus associated with self-reported SSRI response [15]. However, this finding was not replicated in meta-analyses with large-scale population and disorder-focused cohorts [16]. Instead, the meta-analyses revealed one novel locus and estimated the SNP-based heritability at 2%. Although such approaches have yielded valuable insights, it remains unclear whether different measures of SSRI response share a common genetic basis, necessitating further investigation to achieve robust and replicable findings.

This study aims to investigate the genetic architecture of SSRI response by integrating 15 clinical studies comprising 5482 patients with MDD treated with SSRIs. Clinical studies, particularly randomised controlled trials (RCTs), are considered the gold standard for evaluating therapeutic efficacy [17]. Treatment exposure and outcomes are prospectively and systematically assessed using standardised protocols by clinicians. Such rigorous phenotyping minimises ambiguity arising from proxy measures and reduces potential recall or reporting bias. In addition, drug intake is regularly monitored, limiting non-adherence. Although clinical cohorts are modest in sample size for genetic studies, their well-defined phenotypes enable more reliable characterisation of treatment response.

Previous GWAS of antidepressant response have predominantly focused on individuals of European ancestry, leaving gaps in our understanding of the genetic basis of antidepressant response across diverse populations. To address this, we conducted GWAS of clinically assessed SSRI response in European, East Asian, African, and Admixed American ancestries, followed by meta-analyses. We further estimated SNP-based heritability, performed polygenic scoring prediction, and conducted gene-level and gene-set enrichment analyses to advance understanding of common biological underpinnings and inform personalised treatment strategies.

## 2 Methods

### 2.1 Samples and measures

This study included 14 cohorts that shared individual-level genotype data and information on response to treatment, and one cohort that shared GWAS summary statistics, with the PGC MDD Antidepressant Response Working Group. Of these studies, 11 were predominantly of European ancestry and four were of East Asian ancestry. All cohorts were approved by the corresponding ethic boards, and all participants provided written informed consent. All participants had a clinical diagnosis of MDD and were treated with SSRIs. Details of 13 cohorts included in the previous PGC GWAS of antidepressant response [10] are provided in Supplement 1. Two additional studies are included here, the Munich Antidepressant Response Signature (MARS) project and SEOUL_DEP.

Antidepressant response was assessed by two primary outcomes: non-remission and percentage improvement, over a 6-16 week treatment period. Non-remission was determined based on whether patients’ endpoint depression scores remained at or above the clinical cut-off for the rating scale used [18]. Specifically, the pre-defined thresholds for each rating scale were as follows: Montgomery Åsberg Depression Rating Scale (MADRS) > 10, Quick Inventory of Depressive Symptomatology (QIDS) > 5, Hamilton Depression Rating Scale (HAMD-17/HAMD-21) > 7, and Beck Depression Inventory (BDI) > 9 [19–22]. Patients whose scores exceeded these thresholds were classified as non-remitters. Percentage change in depression score from baseline to endpoint was calculated as 100 × (baseline score – endpoint score) / baseline score. This percentage change was then residualised by regressing out age, sex, and baseline depression score using ordinary least squares regression. The resulting residuals were then standardised within each study. The standardised residual score was used as percentage improvement in all subsequent analyses.

#### 2.1.1 MARS

MARS is a prospective naturalistic multi-centre clinical study from Germany [23] (Supplement 1). The project included 1412 European inpatients with a diagnosis of depression recruited from three clinical sites. Treatment and duration of hospitalisation were determined by the patient’s clinical conditions and were not influenced by the study protocol. Antidepressant medications and dosages were monitored and adjusted as per relevant clinical-practice guidelines.

Given the naturalistic study design, patients in the MARS project could receive multiple classes of antidepressants within the same treatment week, and medication switches occurred frequently. To establish a clear definition of SSRI users, the start and end weeks and dosage for each antidepressant received by each patient were extracted, and the duration of use for each drug was calculated. SSRI users were defined as patients who received adequate dosage of SSRIs for at least four weeks, the minimum duration typically required for antidepressants to begin exerting clinical effects (Supplement 1) [24,25].

#### 2.1.2 SEOUL_DEP

The SEOUL_DEP cohort recruited unrelated patients of Korean ancestry diagnosed with MDD from the clinical trials programme of the Samsung Medical Centre (Supplement 1) [26]. Patients received monotherapy with escitalopram, sertraline, fluoxetine, paroxetine, or mirtazapine for 6 weeks. Dose titration was completed within 2 weeks, and clinicians adjusted dose further as necessary. Only participants treated with one of the four SSRIs were included in the current analysis.

### 2.2 Genome-wide association study

#### 2.2.1 Technical/genomic quality control and genotype imputation

Genotyping procedures can be found in original reports of each cohort and are summarised in Supplement 2 S1. GWAS analysis was performed using the Rapid Imputation and COmputational PIpeLIne (RICOPILI) pipeline on Snellius, a high-performance computing (HPC) cluster [27]. The standard workflow is shown in Fig. 1. Details of technical quality control (QC), genomic QC (including principal component analysis (PCA) and relatedness testing), and imputation can be found in Supplement 1.

**Fig. 1:**
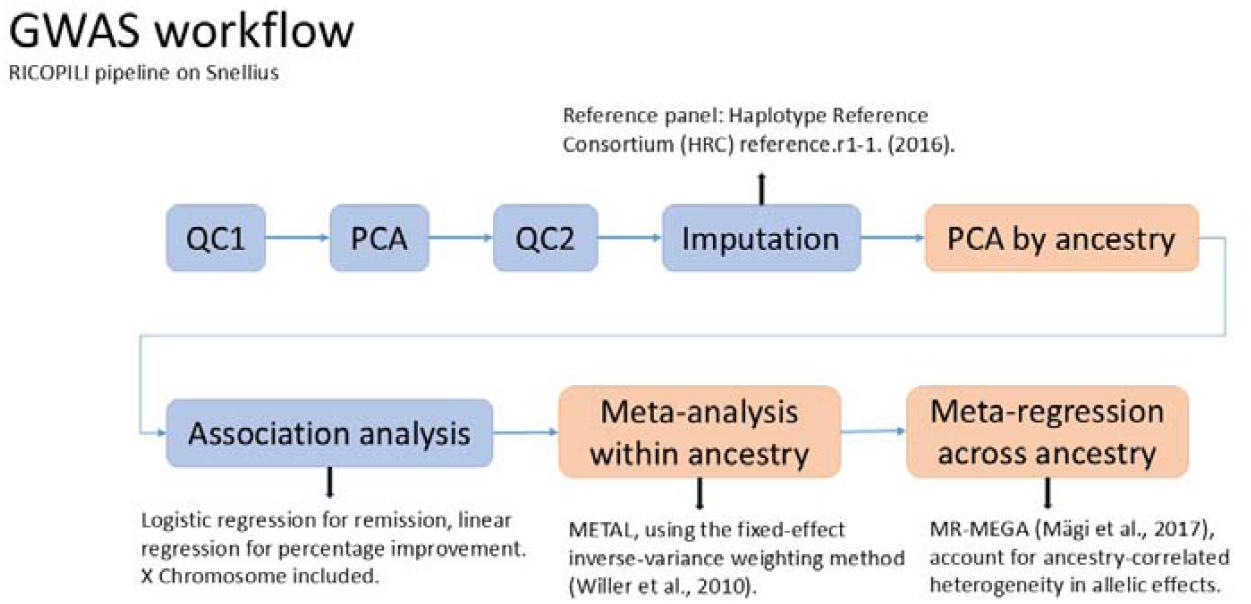
Genome-wide association study (GWAS) workflow. Blue blocks represent steps carried out within individual cohorts, while orange blocks represent steps carried out across cohorts. PCA, principal component analysis; QC, quality control before and after PCA.

Ancestry of the samples was confirmed and assigned by projecting each individual onto principal components derived from the 1000 Genomes reference panel, using EIGENSOFT 6.1.4 [28,29]. Four major ancestry groups were identified: European, East Asian, African, and Admixed American. Analyses were conducted for each ancestry within each study; however, only the STAR*D cohort had an adequate sample size of individuals of African and Admixed American ancestries to permit standalone analyses for these groups.

#### 2.2.2 Association analysis

Association analysis was performed within each cohort, by logistic regression for non-remission and linear regression for percentage improvement. The first four principal components, together with any components nominally significantly associated with non-remission, specifically PC5 for European ancestry, PCs 6 and 12 for East Asian ancestry, and PC20 for African ancestry, were included as covariates in this and subsequent analyses to account for population stratification, along with age, gender, and baseline depression score. For Chromosome X, association analysis was performed in males and females separately, and then meta-analysed by METAL [30]. The effective sample size per case/control group was calculated as N_eff_ = 2 × N_case_ × N_control_ / (N_case_ + N_control_), where N_case_ and N_control_ represent the sample size of non-remitters and remitters, respectively.

#### 2.2.3 Within-ancestry meta-analysis and trans-ancestry meta-regression

Genome-wide summary statistics were then meta-analysed within each ancestry using the inverse-variance weighted fixed-effect meta-analysis in METAL, retaining SNPs with minor allele frequency (MAF) > 0.01 and imputation INFO score > 0.6. Trans-ancestry GWAS meta-regression was performed using MR-MEGA to account for heterogeneity in allele frequencies across ancestries [31]. To identify index variants, Linkage Disequilibrium (LD) clumping was subsequently applied to the MR-MEGA GWAS summary statistics using PLINK v1.9 with the 1000 Genomes Phase 3 reference panel [32]. Clumps were formed around index variants with a p-value smaller than 1×10^−5^. Variants within 1000 kb of an index variant and in LD with it (r^2^ > 0.1) were assigned to the same clump.

### 2.3 SNP-based heritability estimation

SNP-based heritability of non-remission and percentage improvement was estimated using individual-level data from participants of European ancestry with two genomic approaches: genomic-relatedness-based restricted maximum likelihood (GREML) implemented in Genome-wide Complex Trait Analysis (GCTA) [33] and Bayesian multiple regression-based genome-wide complex trait Bayesian analyses (GCTB) [34]. Analyses were restricted to European ancestry cohorts due to the limited sample sizes of other ancestries, and differences in allelic frequencies and LD patterns which precluded meta/mega-analyses across ancestries.

Prior to heritability estimation, cryptic relatedness was addressed in two stages: related individuals were first removed within and across cohorts during GWAS QC in RICOPILI using a PIHAT threshold of > 0.2; followed by a stricter genomic relatedness matrix (GRM) cutoff of 0.05 in GCTA.

Mega-GREML analyses were conducted in GCTA, in which intersected SNPs across studies were extracted and pooled to estimate the heritability jointly, with cohort included as an additional covariate. The SNP-based heritability for non-remission was converted to the liability scale assuming both population and sample prevalence of 0.61, reflecting the proportion of non-remitters across cohorts.

The same set of SNPs and samples were included in a nested BayesS model in GCTB, using non-overlapping 0.2Mb genomic windows, 25,000 Markov chain Monte Carlo iterations and a burn-in of 5,000 iterations. Since heritability estimates can be skewed when the true heritability is close to zero, the posterior mode and 95% highest posterior density (HPD) credible intervals were reported. All estimates were derived after excluding burn-in iterations, and SNP-based heritability for non-remission was likewise converted to the liability scale.

### 2.4 Leave-one-out polygenic scoring prediction

To assess the predictive performance of the GWAS summary statistics for non-remission and percentage improvement, we calculated polygenic scores (PGS) for non-remission and percentage improvement using a leave-one-out design, in which GWAS summary statistics were generated from all cohorts except the target cohort. Polygenic scores were computed using the GenoPred pipeline with SBayesRC [35,36]. SBayesRC integrates GWAS summary statistics with functional genomic annotations and estimates all model parameters internally through a Bayesian Gibbs sampling framework, and was chosen for its strong predictive performance and computational efficiency [37]. Within GenoPred, SNPs are restricted to HapMap3 variants; variants with an INFO score < 0.9, MAF < 0.01 or sample size greater than 3 standard deviations from the median are removed, and LD reference panels matched to the GWAS ancestry are applied. PGS are scaled to the ancestry-matched reference. The resulting PGS were then pooled across cohorts, and mega-regression analyses were conducted to predict the corresponding phenotypic outcomes, including cohort as a covariate.

### 2.5 Shared genetic architecture with psychiatric traits

PGS for 13 psychiatric traits, including MDD [38], anxiety disorders [39], bipolar disorder [40], schizophrenia [41], post-traumatic stress disorder [42], obsessive-compulsive disorder [43], anorexia nervosa [44], attention deficit/hyperactivity disorder (ADHD) [45], autism spectrum disorder (ASD) [46], treatment resistant depression (TRD) [47], SSRI switching defined from electronic health records in UK Biobank [48], SSRI non-response [16], and lithium response for the treatment of bipolar disorder [49] were calculated using the SBayesRC method in the GenoPred pipeline (Supplement 1 Table S1). To avoid sample overlap between MDD GWAS and antidepressant response cohorts in this study, leave-one-out MDD GWAS summary statistics were generated excluding each overlapping cohort (STAR*D, GENetic and clinical Predictors Of treatment response in Depression (GenPod), GENDEP, Group for the Study of Resistant Depression (GSRD), MARS and NEWMEDS (GSK, Pfizer)) from the discovery GWAS used in its corresponding analysis. Associations between PGS and non-remission or percentage improvement were assessed using pooled individual-level data, with logistic and linear regression models, respectively, and cohort as a covariate. A Bonferroni correction was applied to account for multiple testing (13 traits and 2 outcomes tested, α = 0.05/26 = 0.002).

### 2.6 Gene and gene-set analysis

Gene-level association and gene-set enrichment analyses were performed in European samples, using MAGMA v1.10 [50]. Input GWAS summary statistics for non-remission and percentage improvement were subjected to QC filters (MAF > 0.01, INFO > 0.9, sample size per SNP > max(N) × 0.9). Gene-level associations were conducted using the SNP-wise mean model, applying a ±10kb window and LD information from the 1000 Genomes Project phase 3 reference panel. Gene-set analyses were performed using a one-sided competitive test to evaluate whether genes within each set showed greater association with the phenotype than genes outside the set.

As a primary analysis, we tested enrichment in four candidate serotonin-related pathways from KEGG: serotonergic synapse (hsa04726), neuroactive ligand signalling (hsa04082), tryptophan metabolism (hsa00380), and folate biosynthesis (hsa00790) [51,52].

To investigate the clinical relevance of identified genetic signals, drug enrichment analyses were performed using drug-gene sets derived from Drug Targetor (1761 drug-gene sets; wholedatabase_for_targetor) [53]. Drug-gene sets were then grouped into Anatomical Therapeutic Chemical (ATC) classes at three hierarchical levels: therapeutic subgroup (2nd level), pharmacological subgroup (3rd level), and chemical subgroup (4th level). For each ATC class, drug class enrichment was assessed by calculating the area under the curve (AUC) of the ranked drug-level p-values using the Wilcoxon rank-sum test, where AUC > 0.5 indicates that drugs within the class tend to show stronger association with the phenotype than drugs outside the class.

We then expanded analyses across gene-sets from the Human Molecular Signatures Database (MSigDB) Collections (35134 gene sets; msigdb.v2025.1.Hs.entrez.gmt) [54,55].

Bonferroni correction was applied to account for multiple testing: 18,395 gene-level tests (α = 2.72 × 10^−6^); 4 serotonin-related pathways (α = 0.0125); 35,086 MSigDB gene-sets (α = 1.43 × 10^−6^); 1463 drug-gene sets in Drug Targetor (α = 3.42 × 10^−5^); 36,553 combined genel□set analyses (stringent α = 1.37 × 10^−6^); and 610 drug-class enrichment tests (α = 8.20 × 10^−5^). If any gene-set was significantly enriched after Bonferroni correction, the constituent genes were identified from the respective database, and MAGMA gene-level p-values were retrieved for each gene to determine which gene most strongly drove the enrichment signal.

### 2.7 Statistical analyses

All statistical analyses were conducted on the Snellius HPC using R (version 4.3.2) [56].

### 2.8 Sensitivity analyses

Sensitivity analyses, including GWAS, SNP-based heritability estimation, and leave-one-out polygenic scoring prediction, were repeated in European-ancestry cohorts treated exclusively with SSRIs, to assess the robustness of findings in a more homogeneous subset. Seven studies were analysed, after excluding three naturalistic studies (Depression and Sequence of Treatment (DAST), MARS, and GSRD) and GODS due to frequent drug switching.

## 3 Results

### 3.1 Study overview

A total of 5502 participants were identified as SSRI users from the 15 cohorts included. Only the STAR*D cohort had sufficient participants of African or Admixed American ancestry for analysis (with sample sizes of <20 in other studies). After quality control, 5482 participants with information on percentage improvement and 5467 participants with non-remission data were retained (Table 1; Supplement 1), of whom 61.5% were classified as non-remitters (Fig. 2), ranging from 35.0% - 81.1% of participants in each cohort.

**Table 1.** Summary of SSRI users in 15 studies analysed, by ancestry analysis group.

| Cohort name | Initiated country/region | Study design | Primary measure | Medications | Age (years), mean (SD) | Sex (female), N (%) | Baseline severe cases, % | Baseline depression score, mean (SD) | No. of remitters | No. of non-remitters (%) | No. of percentage improvement |
| --- | --- | --- | --- | --- | --- | --- | --- | --- | --- | --- | --- |
| European ancestry |  |  |  |  |  |  |  |  |  |  |  |
| DAST | Germany | Naturalistic | HAMD-21 | Various | 46.6 (17.3) | 80 (66.1) | 40.5 | 21.8 (7.9) | 56 | 65 (53.7) | 121 |
| GENDEP | 9 EUR countries | Partial RCT | MADRS | Escitalopram | 42.4 (11.5) | 276 (62.2) | 16.9 | 28.4 (6.7) | 200 | 230 (53.5) | 444 |
| GENPOD | United Kingdom | RCT | BDIs | Citalopram | 39.3 (12.1) | 158 (67.0) | 69.9 | 34.0 (9.4) | 87 | 149 (63.1) | 236 |
| GODS | Switzerland | Partial RCT | MADRS | Paroxetine | 37.1 (10.3) | 39 (53.4) | 28.8 | 31.8 (4.7) | 17 | 56 (76.7) | 73 |
| GSRD | 8 EUR countries | Naturalistic | MADRS | Various | 52.2 (14.3) | 379 (68.0) | 39.9 | 33.4 (7.3) | 105 | 452 (81.2) | 557 |
| MARS | Germany | Naturalistic | HAMD-21 | Various | 46.7 (14.5) | 123 (61.5) | 46.5 | 22.6 (7.4) | 93 | 101 (52.1) | 194 |
| MAYO | United States | Open label | HAMD-17 | Citalopram, Escitalopram | 40.0 (13.9) | 96 (61.9) | 26.9 | 21.0 (5.0) | 80 | 75 (48.4) | 156 |
| NEWMEDS (GSK) | United States | RCT | HAMD-17 | Escitalopram | 36.4 (11.9) | 72 (54.6) | 38.6 | 23.0 (3.1) | 56 | 76 (57.6) | 132 |
| NEWMEDS (Pfizer) | United States | RCT | HAMD-17 | Fluoxetine, Paroxetine, Sertraline | 43.2 (13.1) | 208 (67.3) | 48.2 | 23.5 (3.4) | 99 | 210 (68.0) | 309 |
| PGRN-AMPS | United States | Open label | QIDS-C<sub>16</sub> | Citalopram, Escitalopram | 39.9 (13.7) | 306 (62.2) | 44.8 | 15.1 (3.5) | 199 | 292 (59.5) | 491 |
| STAR*D | United States | Single arm | QIDS-SR<sub>16</sub> | Citalopram | 43.3 (13.5) | 678 (57.8) | 55.5 | 16.2 (3.2) | 506 | 668 (56.9) | 1174 |
| EUR total |  |  |  |  |  |  |  |  | 1498 | 2374 (61.3) | 3887 |
| East Asian ancestry |  |  |  |  |  |  |  |  |  |  |  |
| Japan | Japan | RCT | HAMD-17 | Fluvoxamine, Paroxetine | 46.0 (15.3) | 56 (46.7) | 26.7 | 20.1 (5.7) | 77 | 40 (34.2) | 117 |
| Miaoli | Taiwan | Open label | HAMD-17 | Escitalopram, Paroxetine | 41.4 (13.6) | 192 (82.4) | 30.0 | 21.3 (4.2) | 102 | 125 (55.1) | 227 |
| Taipei | Taiwan | Open label | HAMD-17 | Citalopram, Fluoxetine, Sertraline | 47.0 (15.1) | 96 (55.2) | 79.3 | 26.5 (4.1) | 43 | 126 (74.6) | 169 |
| SEOUL_DEP | Korea | Open label | HAMD-17 | Various | 60.3 (13.7) | 404 (72.8) | 13.5 | 20.1 (4.1) | 202 | 353 (63.6) | 555 |
| EAS total |  |  |  |  |  |  |  |  | 424 | 644 (60.3) | 1068 |
| African ancestry |  |  |  |  |  |  |  |  |  |  |  |
| STAR*D | United States | Single arm | QIDS-SR<sub>16</sub> | Citalopram | 44.3 (12.6) | 172 (62.1) | 62.8 | 17.1 (3.5) | 93 | 184 (66.4) | 277 |
| Admixed American ancestry |  |  |  |  |  |  |  |  |  |  |  |
| STAR*D | United States | Single arm | QIDS-SR<sub>16</sub> | Citalopram | 41.0 (13.4) | 176 (70.4) | 64.8 | 16.7 (3.1) | 92 | 158 (63.2) | 250 |
| Total |  |  |  |  |  |  |  |  | 2107 | 3360 (61.5) | 5482 |

DAST, Depression and Sequence of Treatment; GENDEP, Genome Based Therapeutic Drugs for Depression; GENPOD, GENetic and clinical Predictors Of treatment response in Depression; GODS, Geneva Outpatient Depression Study; GSRD, Group for the Study of Resistant Depression; MARS, Munich Antidepressant Response Signature project; NEWMEDS, Novel methods leading to new medications in depression and schizophrenia; GSK, Glaxo Smith Kline; PGRN-AMPS, Pharmacogenomics Research Network Antidepressant Medication Pharmacogenomic Study; STAR\*D, Sequenced Treatment Alternatives to Relieve Depression; RCT, randomised controlled trials; HAMD-17, 17-item Hamilton Depression Rating Scale; HAMD-21, 21-item Hamilton Depression Rating Scale; MADRS, Montgomery Åsberg Depression Rating Scale; BDI, Beck Depression Inventory; QIDS-C<sub>16</sub>, 16-item Quick Inventory of Depressive Symptomatology Clinician Rating version; QIDS-SR<sub>16</sub>, 16-item Quick Inventory of Depressive Symptomatology Self-Report version.

**Fig. 2:**
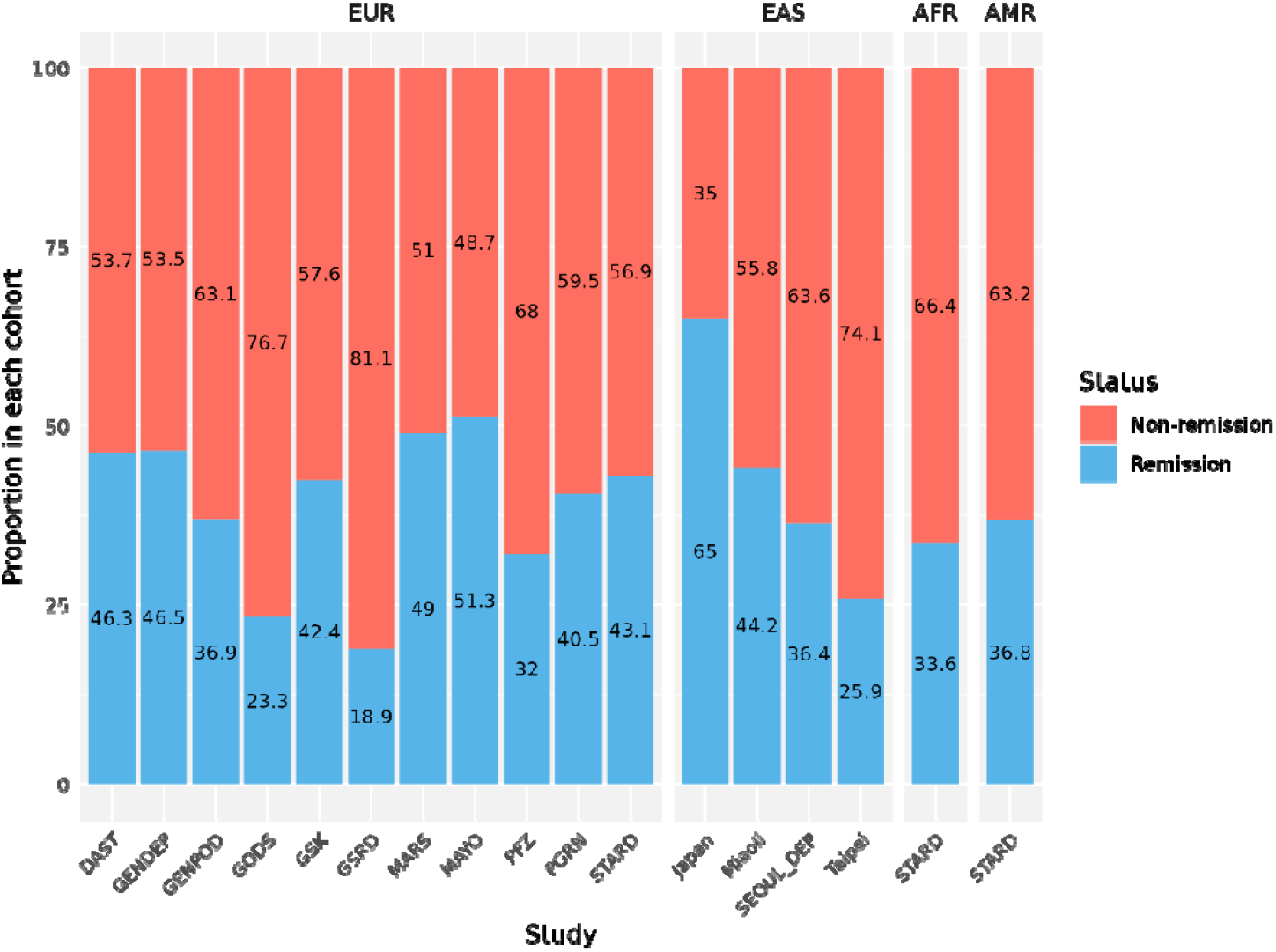
Proportion of participants remaining non-remission in 15 cohorts, by ancestry. Across all studies, 61.5% of participants did not attain remission. EUR: European; EAS: East Asian; AFR: African; AMR: Admixed American.

### 3.2 Genome-wide analysis

For each outcome, we carried out four ancestry-specific GWAS and one trans-ancestry GWAS meta-regression. Specifically, for European ancestry, 2374 non-remitters and 1498 remitters were included (N_eff_: 1766); for East Asian ancestry, we combined GWAS summary statistics derived from individual-level genotypes from 3 cohorts with summary statistics shared by the SEOUL_DEP cohort, resulting in 644 non-remitters and 424 remitters (N_eff_: 485); for African ancestry, 184 non-remitters and 93 controls were included (N_eff_: 123); for Admixed American ancestry, 158 non-remitters and 92 remitters were included (N_eff_: 116). Additional details are provided in Supplement 2 S1.

No variants were associated with non-remission or percentage improvement at the genome-wide significant level in any ancestry, or in the trans-ancestry meta-regression, as illustrated in the Manhattan plots (Fig. 3) and QQ-plots (Supplement 1 Figure S5). Trans-ancestry meta-regression yielded 8 independent loci with suggestive associations (p < 1 × 10^−5^) for non-remission, and 17 loci for percentage improvement (Supplement 2 S2-S11). The strongest signal for non-remission was rs7753048 on chromosome 6 (p = 1.02 × 10^−7^), and for percentage improvement was rs6984323 on chromosome 8 (p = 9.82 × 10^−8^).

**Fig. 3:**
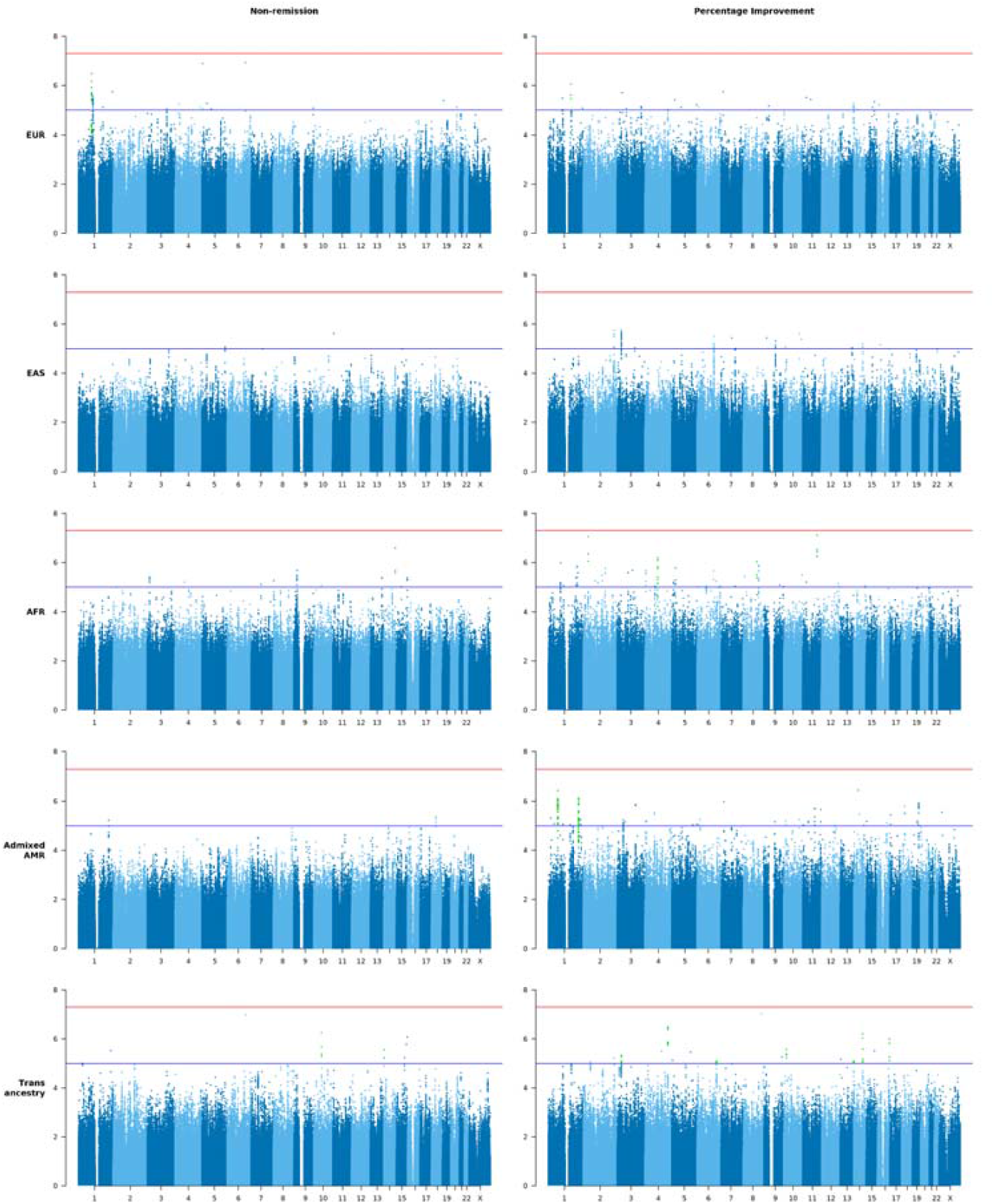
Manhattan plots for GWAS of non-remission (left) and percentage improvement (right) across four ancestries and trans-ancestry meta-regression. Horizontal lines indicate genome-wide significance (p = 5 × 10^−8^; red) and suggestive significance (p = 1 × 10^−5^; blue). Green points indicate lead SNPs supported by multiple variants in linkage disequilibrium (r^2^ > 0.6), with INFO > 0.6.

### 3.3 SNP-based heritability estimation

After removing individuals with cryptic relatedness, 3820 participants of European ancestry were included in the heritability estimation. A total of 1,522,029 intersected SNPs were analysed. Mega-GREML analyses did not identify significant evidence for non-zero heritability for either non-remission or percentage improvement. The mega-GREML estimate of SNP-based heritability for non-remission was *h*^2^ = 0.118 (SE = 0.082, 95% CI [-0.043, 0.279], p = 0.070, assuming a liability scale population and sample prevalence of 0.61), and for percentage improvement was *h*^2^ = 0.000 (SE = 0.049, 95% CI [-0.097, 0.096], p = 0.497). Using the nested BayesS model in GCTB, SNP-based heritability for non-remission was estimated at a posterior mode of 0.046 (95% HPD credible intervals [0.010, 0.171], and for percentage improvement at a posterior mode of 0.030 (95% HPD credible intervals [0.007, 0.098]), assuming both population and sample prevalence of 0.61; Fig. 4).

**Fig. 4:**
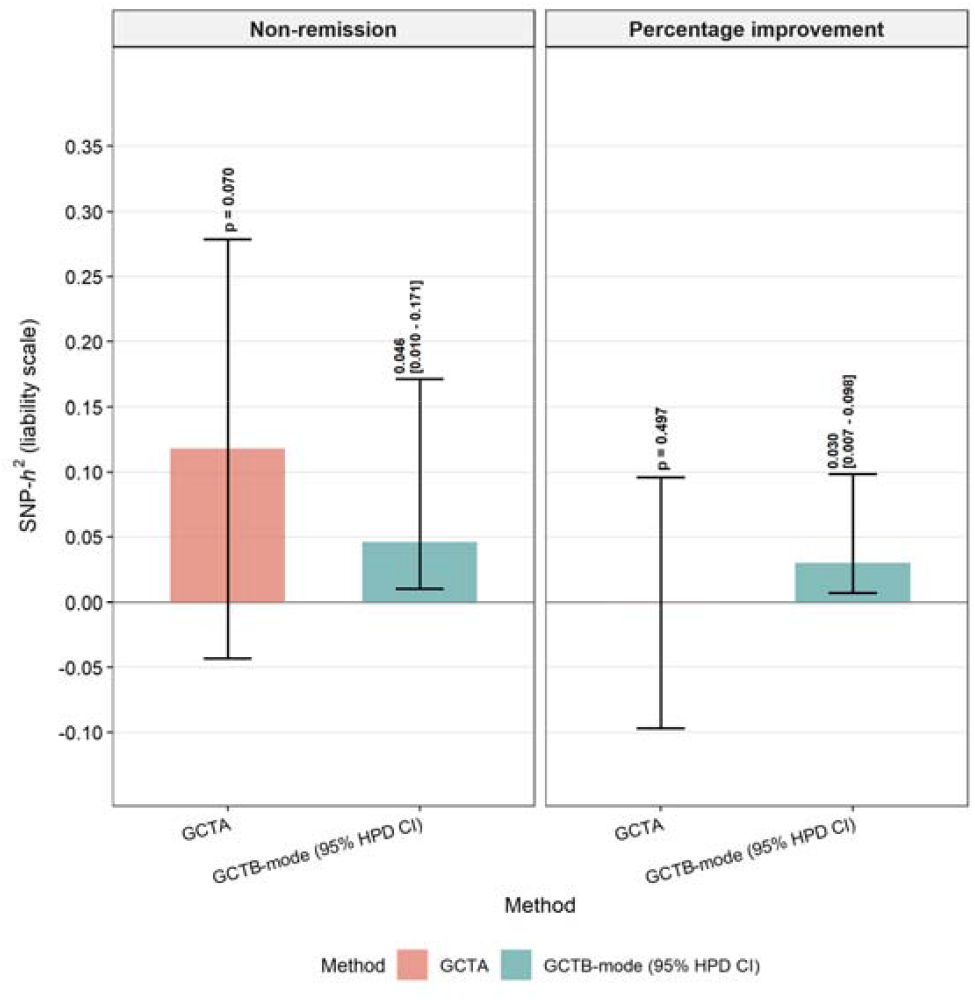
Single Nucleotide Polymorphism-based heritability estimation for non-remission and percentage improvement by GCTA and GCTB. Estimates for non-remission were converted to a liability scale assuming both population and sample prevalence of 0.61. GCTA, Genome-wide Complex Trait Analysis; GCTB, genome-wide complex trait Bayesian analyses; HPD, highest posterior probability.

### 3.4 Polygenic scoring prediction

We conducted leave-one-out GWAS meta-analysis for non-remission and for percentage improvement, then calculated PGS to evaluate their predictive performance. Neither leave-one-out polygenic scores for non-remission (OR = 1.007, 95% CI [0.941, 1.078], p = 0.837) or percentage improvement (β = -0.017, 95% CI [-0.050, 0.016], p = 0.310) were significantly associated with their matched phenotypes (Fig. 5).

**Fig. 5:**
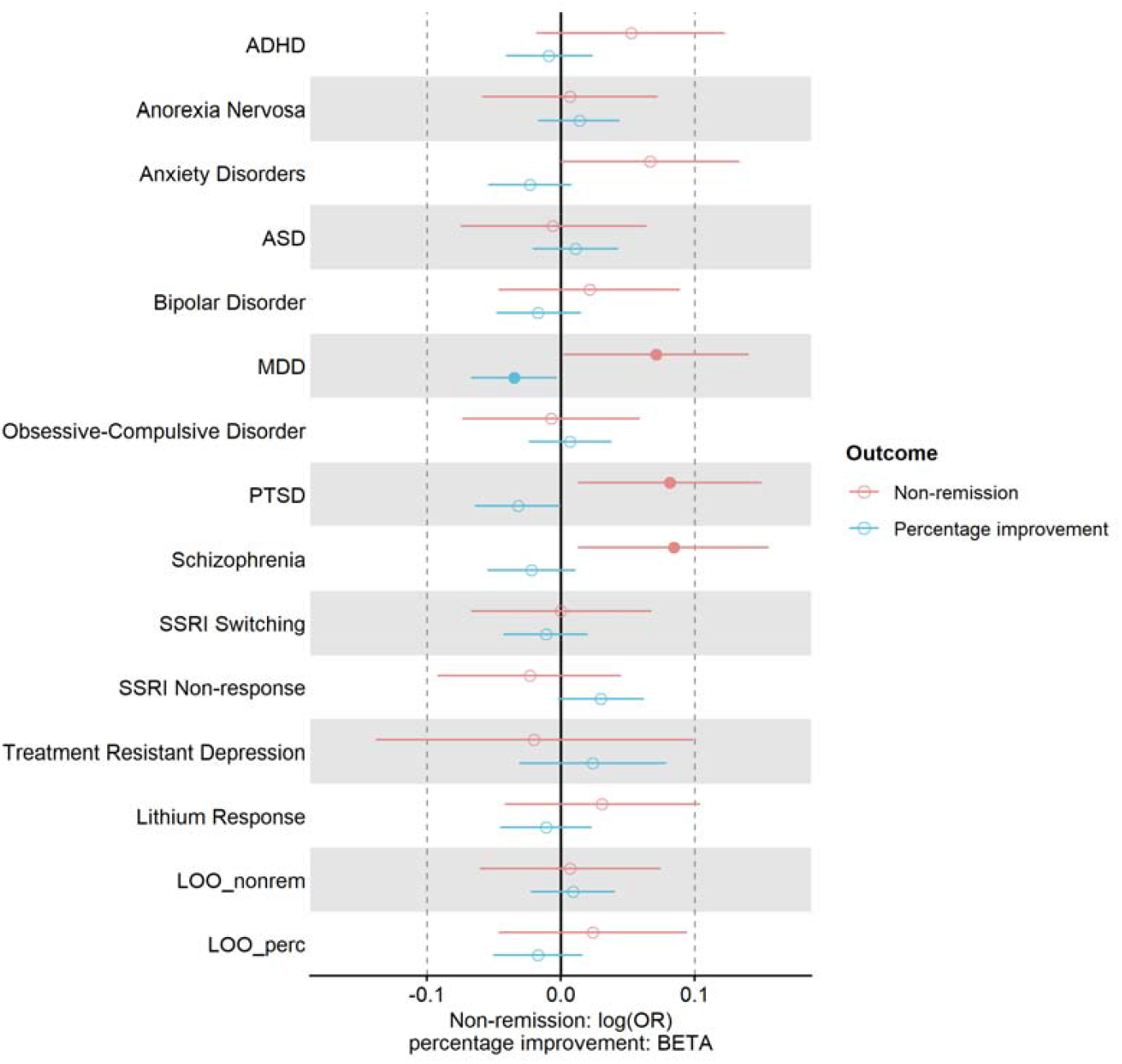
Shared genetic architecture between SSRI response and 13 psychiatric traits, and leave-one-out prediction. Associations between psychiatric trait polygenic scores and antidepressant response outcomes were assessed to investigate shared genetic architecture. ADHD, attention deficit/hyperactivity disorder; ASD, autism spectrum disorder; MDD, major depressive disorder; PTSD, post-traumatic stress disorder; LOO_nonrem, leave-one-out PGS for non-remission; LOO_perc, leave-one-out PGS for percentage improvement.

We tested whether polygenic scores for psychiatric traits were associated with antidepressant response. Higher non-remission rates were nominally associated with higher PGS for MDD (OR = 1.074, 95% CI [1.002, 1.151], p = 0.042), PTSD (OR = 1.085, 95% CI [1.013, 1.168], p = 0.021), and schizophrenia (OR= 1.088, 95% CI [1.013, 1.168], p = 0.020), while better percentage improvement was associated with lower PGS for MDD (β = -0.035, 95% CI [-0.067, -0.003], p = 0.032), as illustrated in Fig. 5 and Supplement 1 Table S2. However, none of these associations survived multiple testing correction (p < 0.002).

### 3.5 Gene and gene-set analysis

Gene-level association analyses did not identify any signal reaching the threshold for genome-wide significance (p < 2.72 × 10^−6^) for either non-remission or percentage improvement (Supplement 2 S12-S13). The strongest associations showed consistency across outcomes: *FXYD5* on chromosome 19 (non-remission p = 8.38 × 10^−5^; percentage improvement p = 1.08 × 10^−4^), *DHX8* on chromosome 17 (non-remission p = 9.80 × 10^−5^; percentage improvement p = 2.77 × 10^−4^), and *ETV4* on chromosome 17 (non-remission p = 1.16 × 10^−4^; percentage improvement p = 1.03 × 10^−4^), which were in the top 5 signals in both outcomes. Additional top signals for non-remission were observed at *TMEM56-RWDD3/TMEM56* on chromosome 1 (p = 4.44 × 10^−5^ and p = 7.04 × 10^−5^, respectively), and at *PROP1* on chromosome 5 (p = 1.07 × 10^−4^) for percentage improvement.

Gene-set enrichment analyses revealed that among the four serotonin-related KEGG pathways, genes associated with non-remission were significantly enriched in the serotonergic synapse pathway (N_genes = 105, β = 0.197, SE = 0.086, p = 0.011). None of the MSigDB or Drug Targetor gene sets reached genome-wide significance (p < 1.43 × 10^−6^ and p < 3.42 × 10^−5^, respectively; Supplement 2 S14-S19, Supplement 1 Figure S8-S11). Drug-class enrichment analyses identified significant associations for quinolone antibacterials with both non-remission and percentage improvement (ATC code J01M; non-remission: AUC = 0.872, p = 4.82 × 10^−9^; percentage improvement: AUC = 0.838, p = 9.58 × 10^−8^). Atypical antipsychotics were significantly associated with percentage improvement (ATC code N05AX, AUC = 0.929, p = 4.44 × 10^−5^), whereas antimigraine preparations were associated with non-remission (ATC code N02C, AUC = 0.835, p = 1.50 × 10^−5^; Supplement 2 S20-S21).

### 3.6 Sensitivity analyses

Seven European ancestry cohorts (GENDEP, GenPod, Mayo, NEWMEDS (GSK, Pfizer), PGRN-AMPS, and STAR*D) were included in the sensitivity analyses. GWAS of non-remission or percentage improvement did not identify any genome-wide significant variants. SNP-based heritability estimation (N = 2900) did not detect significant heritability for either non-remission (observed scale *h*^2^ = 0.097, SE = 0.081; converting to a liability scale assuming a population prevalence of 0.61 and sample prevalence of 0.58, *h*^2^ = 0.153, SE = 0.128, p = 0.113) or percentage improvement (*h*^2^ = 0.096, SE = 0.081, p = 0.117) in GCTA. In addition, leave-one-out polygenic scores for percentage improvement or non-remission were not significantly associated with their corresponding phenotypes.

## 4 Discussion

We present the largest trans-ancestry GWAS of response to SSRIs in a meta-analysis of 15 clinical studies with 5482 participants diagnosed with MDD. No genome-wide associations were identified for non-remission or percentage improvement in any single ancestry group or in the trans-ancestry meta-regression. This lack of detectable signals may reflect limited statistical power – the effective sample size was only 2490 participants per case/control group even in the trans-ancestry GWAS meta-regression. In the trans-ancestry meta-regression, 8 independent loci showed tentative evidence of association with non-remission and 17 loci with percentage improvement, reaching a significance threshold of p < 1 × 10^−5^.

Genes associated with non-remission or percentage improvement at 1×10^−4^ level included those involved in neurodevelopmental regulation, energy dependency, and lipid homeostasis. Notably, genes *ETV4* and *DHX8* were identified as significantly associated in our previous, partially overlapping GWAS of antidepressant response across all drug classes [10]. *FXYD5* is the subunit of the Na, K-ATPase sodium-potassium pump [57], potentially serving as the energy dependency of serotonin transporter. *TMEM56* (also known as *TLCD4*) is a transmembrane protein, predicted to be involved in lipid homeostasis [58].

We did not detect any significant SNP- or gene-level associations for cytochrome P450 enzymes involved in SSRI metabolism, including *CYP2D6* (non-remission p = 0.335, percentage improvement p = 0.281) and *CYP2C19* (non-remission p = 0.056, percentage improvement p = 0.786) in either GWAS or MAGMA gene-level analyses. Any true effects are likely diluted because these enzymes have drug-specific metabolic profiles, while our analysis combined individuals taking different SSRIs. Additionally, *CYP2D6* structural variants cannot be reliably imputed from SNP-array genotype data. In a previous analysis of imputed *CYP2C19* metabolising status in these studies, evidence for association with antidepressant response was limited, despite accurate imputation from genotype data [59].

Pathway enrichment analyses showed that genes associated with non-remission were significantly enriched in the serotonergic synapse pathway, which is involved in the networks of serotonin metabolism, transport, and serotonin receptor signalling [60]. This enrichment was primarily driven by genes involved in transport of serotonin, including *SLC6A4*, which showed nominal association with non-remission (64 SNPs annotated to the gene; p = 0.034). However, this pathway did not reach significance after multiple testing correction in the broader analysis of pathways from MSigDB.

No significant signal for SNP-based heritability was identified for either non-remission or percentage improvement among participants of European ancestry using GCTA. The mega-GREML estimate for non-remission was 12%, but its confidence interval included zero. While GCTB yielded non-zero 95% HPD credible intervals for both phenotypes, this likely reflects the right-skewed posterior distribution expected under limited sample size rather than strong evidence of heritability. The posterior mode was low for both phenotypes, suggesting that any underlying genetic contribution is likely modest. With only 3820 participants, the study had only 16% power to detect a non-zero heritability [61]. When analyses were restricted to pure SSRI cohorts and naturalistic studies were excluded, heritability estimates converged across both phenotypes at approximately 9% (p ≈ 0.1 for both), suggesting that inclusion of naturalistic studies introduced heterogeneity that differentially affected the two phenotypes. The similarity of estimates in the restricted sample is noteworthy, as it implies a comparable genetic contribution to both non-remission status and symptomatic improvement following SSRI treatment, despite insufficient power to reach significance in the current sample. In contrast, using a substantially larger sample (N = 114,324), Koch et al. [16] detected significant SNP-based heritability for SSRI non-response (*h*^*2*^ = 0.020, SE = 0.006, p < 0.001) using Linkage Disequilibrium Score Regression (LDSC) in a GWAS meta-analysis. While their study benefited from larger sample size, it had more heterogeneous measures of SSRI response, primarily derived from self-reported questionnaires and SSRI switching patterns in electronic healthcare records, differing significantly from the stringent clinically defined non-remission phenotype used in this study.

Polygenic score analyses revealed nominal associations between SSRI response and psychiatric traits, none of which survived multiple testing correction. Better response was nominally associated with lower genetic predispositions for MDD, PTSD, and schizophrenia. Notably, the higher PGS for schizophrenia among non-remitters is consistent with previous findings across antidepressant classes [10], as well as reports of associations with self-reported non-response to SSRIs [62], and TRD [63]. This pattern may reflect shared genetic pathways or clinical trajectories, whereby individuals with depression have an increased risk of developing schizophrenia subsequently [64]. Further evidence comes from the drug class enrichment analysis, where atypical antipsychotics were significantly associated with percentage improvement. This genetic observation aligns with clinical efficacy of atypical antipsychotic augmentation in MDD and TRD [65–67].

Higher PGS for MDD were observed among non-remitters despite covarying for baseline severity. This finding may reflect depressive symptoms being more likely to persist in individuals with higher genetic liability for MDD, thereby reducing the likelihood of antidepressant effectiveness in the short time period of clinical trials. Higher genetic susceptibility to PTSD was also observed among non-remitters. However, this may reflect shared genetic architecture and high interconnected symptoms with depression rather than independent effects [68,69].

Two complementary measurements of SSRI response were used in this study. Non-remission has high clinical utility and is defined using a stringent cut-off on depression rating scales, while percentage improvement measures change in depression symptoms after treatment as a quantitative outcome [70], and may capture more information. For example, an individual with severe depression and high baseline symptom severity may not reach remission after 6-12 weeks of treatment, but may show substantial percentage improvement in their depressive symptoms. The phenotypic correlation between non-remission and percentage improvement is -0.72, indicating strong consistency between the two measurements of SSRI response. However, genetic analysis of non-remission showed better powered results, in heritability estimation, polygenic scoring prediction, gene-level analysis, and gene-set enrichment analysis. Percentage improvement, by contrast, may introduce greater random variation, especially given the cohort heterogeneity which may remain after adjustment for age, gender, and baseline score, and standardisation. For instance, the cohorts employed four different rating scales, each with distinct emphases [71]. HAMD scales place greater weights on somatic symptoms, including three items assessing sleep disturbances alone, whereas BDI focuses more on cognitive and affective symptoms, with only one item for sleep disturbance. Consequently, improvement within a specific symptom domain may translate into different percentage improvement depending on the scale used. These considerations suggest that future studies may benefit from analysing symptom-specific changes rather than total scores, as well as from tracing longitudinal trajectories of antidepressant response.

This study has several limitations. Although 15 cohorts with prospectively assessed treatment response were analysed, the sample sizes were limited for genetic analyses. We specifically focused on SSRIs to determine whether the genetics underpinnings of response in a single drug class would be more tractable than combining all classes of antidepressants. However, no genome-wide significant associations were identified and SNP-based heritability estimates were not different from zero. This contrasts with the significant heritability estimation in our previous analysis of all antidepressant classes [10], although the estimates of non-remission are similar at 0.12. PGS predictions of response across studies did not yield significant results, indicating a lack of statistical power in this study. This research is also limited by heterogeneity within study cohorts, such as symptom differences, and across cohorts. Three cohorts employed naturalistic study design, bringing in effects from other classes of antidepressants on top of SSRIs. More homogeneous study designs, ideally randomised controlled trials with placebo arms, would be valuable for isolating SSRI-specific genetic factors.

This study provides the first trans-ancestry meta-regression of SSRI response, with analyses performed for participants of East Asian, African, and Admixed American ancestries, but few studies were available, and African and Admixed American ancestry groups were represented solely by the STAR*D cohort. More cohorts representing diverse ancestries are therefore necessary.

In summary, larger cohorts will be required to identify genetic contributions to antidepressant response—across all drugs, by class, and by individual drug. While clinical trials remain the gold standard for assessing treatment effects, incorporating real-world data such as drug-switching, or patient-reported outcomes will be valuable for disentangling the highly polygenic architecture underlying antidepressant response. These efforts will be greatly facilitated by wide sharing of diverse ancestry summary statistics from this study and others.

## Supporting information

supplement documenting cohort information, additional methods and result figures.

supplementary tables for results from genome-wide association studies, gene and gene-set enrichment analysis.

## Data Availability

All data produced in the present study are available upon reasonable request to the authors.

## Acknowledgements

This work was supported by the National Institutes of Health (MH124873 and MH124871), the Wellcome Trust [226770/Z/22/Z; 222811/Z/21/Z] and the National Institute for Health and Care Research (NIHR) Maudsley Biomedical Research Centre (BRC). We thank SURF (www.surf.nl) for the support in using the National Supercomputer Snellius. KH is grateful for the financial support provided by the Program of China Scholarship Council (Grant No. 202408060066).

We gratefully acknowledge the participants and collaborators who contributed to the data. The collection of the sample from the Group for the Study of Resistant Depression (GSRD) Consortium was supported by an unrestricted grant from Lundbeck for the GSRD. Lundbeck had no further role in the study design and the collection, analysis, and interpretation of data. The GENDEP (Genome Based Therapeutic Drugs for Depression) study was funded by a European Commission Framework 6 grant (EC Contract Ref. No. LSHB-CT-2003–503428). H. Lundbeck provided nortriptyline and escitalopram for the GENDEP study. GlaxoSmithKline and the UK National Institute for Health Research of the Department of Health contributed to the funding of the sample collection at the Institute of Psychiatry, London. GENDEP Illumina array genotyping was funded in part by a joint grant from the UK Medical Research Council and GlaxoSmithKline (Grant No. G0701420). The GENPOD (GENetic and clinical Predictors Of treatment response in Depression) trial was funded by the UK Medical Research Council and supported by the UK Mental Health Research Network. The genotyping of GENPOD samples was supported by the Innovative Medicine Initiative Joint Undertaking under Grant No. 115008, of which resources are composed of European Union and the European Federation of Pharmaceutical Industries and Associations (EFPIA) in-kind contribution and financial contribution from the European Union’s Seventh Framework Programme (Grant No. FP7/2007–2013). EFPIA members Pfizer, GlaxoSmithKline, and F. Hoffmann-La Roche have contributed work and samples to the project presented here. The PGRN-AMPS (Pharmacogenomics Research Network Antidepressant Medication Pharmacogenomic Study) study data were obtained via the database of Genotypes and Phenotypes (dbGAP) (Accession No. phs000670.v1.p1). Funding support for the PGRN-AMPS study was provided by the National Institute of General Medical Sciences, National Institutes of Health, through the PGRN grant to Principal Investigators R. Weinshilboum and L. Wang (Grant No. U19 GM61388). D. Mrazek served as the Principal Investigator for the PGRN-AMPS study within the Mayo Clinic PGRN program. Genome-wide genotyping was performed at the RIKEN Center for Genomic Medicine, with funding provided by RIKEN. The datasets used for the analyses described in this manuscript were obtained from the dbGaP at http://www.ncbi.nlm.nih.gov/gap/. STAR*D genotyping was led by SP Hamilton and funded by National Institute of Mental Health (NIMH; Award No. R01 MH-072802). SEOUL_DEP was supported by grants from the Basic Science Research Program through the National Research Foundation of Korea (NRF) funded by the Ministry of Education, Science and Technology (NRF-2014R1A2A1A10052419, to DKK), the Korean Health Technology R&D Project through the Korea Health Industry Development Institute (KHIDI) funded by the Ministry of Health & Welfare, Republic of Korea (HI14C2071, to DDK), Lundbeck (to DDK) and the Brain Research Program through the National Research Foundation of Korea (NRF) funded by the Ministry of Science, ICT & Future Planning (NRF-2014M3C7A1046049, to J-WK). The NEWMEDS study was funded by the Innovative Medicine Initiative Joint Undertaking (IMI-JU) under grant agreement no. 115008 of which resources are composed of European Union and the European Federation of Pharmaceutical Industries and Associations (EFPIA) in-kind contribution and financial contribution from the European Union’s Seventh Framework Programme (FP7/2007-2013).

