## supplement documenting cohort information, additional methods and result figures. for "Trans-ancestry genome-wide association meta-analysis of antidepressant response to selective serotonin reuptake inhibitors in clinical studies of depression"

**Hu et al.**

Supplementary materials

Table of Contents

1 Cohort information	2

2 Measures of antidepressant response	10

3 Descriptive statistics across cohorts	11

4 Quality control and imputation using RICOPILI	12

5 QQ-plots for GWAS	13

6 SNP-based heritability estimation	16

7 Shared genetic architecture with 13 psychiatric traits	16

8 Gene-set level analyses	19

PGC MDD Working Group authorship list……………………………………………………………………….25

References……………………………………………………………………………………………………………………..35

### 1 Cohort information

**European ancestry**

MARS

MARS is a prospective naturalistic multi-centre clinical study initiated by the Max Planck Institute of Psychiatry in Germany, aiming to identify predictors for drug response and stratify homogeneous subtypes of depression [1]. The project included 1412 inpatients with a diagnosis of depression from three clinical sites in southern Bavaria. All participants were of European ancestry, aged between 18 and 75 years, and diagnosed with a single major depressive episode, recurrent depression, or bipolar depression by trained psychiatrists in accordance with the DSM-IV criteria. Patients experiencing depressive syndromes secondary to medical or neurological conditions, manic, hypomanic, or mixed affective symptoms, alcohol dependence, drug abuse, or severe physical medical conditions were excluded.

The study was approved by the local Ethics Committee of the Ludwig Maximilians University, Munich, Germany, and carried out in accordance with the latest version of the Declaration of Helsinki.

The project collected sociodemographic and clinical information, including age at onset of first MDD episode, number of previous depressive episodes, and family history of psychiatric disorders at admission. Treatment and duration of hospitalisation were determined based on the patient’s clinical condition and were not influenced by the study protocol. Antidepressant medications and dosages were monitored and adjusted, with plasma concentrations assessed weekly from admission to week 4, then bi-weekly until discharge (up to a maximum of 16 weeks). Depression severity was primarily measured using the 21-item Hamilton Depression Rating Scale (HAMD-21) on the same schedule.

Due to the naturalistic study design, patients in the MARS project could have received multiple classes of antidepressants within the same treatment week, and medication switches occurred frequently, so it was essential to clarify the definition of SSRI users. We first excluded participants who did not receive any common antidepressant medications (including SSRIs, SNRIs, TCAs, SSRE (tianeptine), NaSSA (mirtazapine), NaRI (reboxetine), and MAOI), resulting in a sample of 1173 participants. Next, we extracted the start and end weeks, as well as the dosage, for each antidepressant received by each patient, and calculated the duration of use for each drug. We defined SSRI users as patients who received adequate dosages of SSRIs for at least four weeks, which is the minimum duration typically required for antidepressants to begin showing effects [2]. The usual minimum effective dosages for each SSRIs were as follows: escitalopram ≥ 10mg/day, citalopram ≥ 20mg/day, paroxetine ≥ 20mg/day, sertraline ≥ 50mg/day, fluoxetine ≥ 20mg/day, vortioxetine ≥ 10mg/day, vilazodone ≥ 10mg/day [3].

In the MARS project, the baseline was defined as the week a patient started receiving an SSRI, rather than automatically using the admission week as in other cohorts. The endpoint was defined as the week the patient stopped that SSRI treatment, rather than prior to discharge. Non-remission and percentage improvement were calculated based on this period of treatment, standardised among SSRI users only.

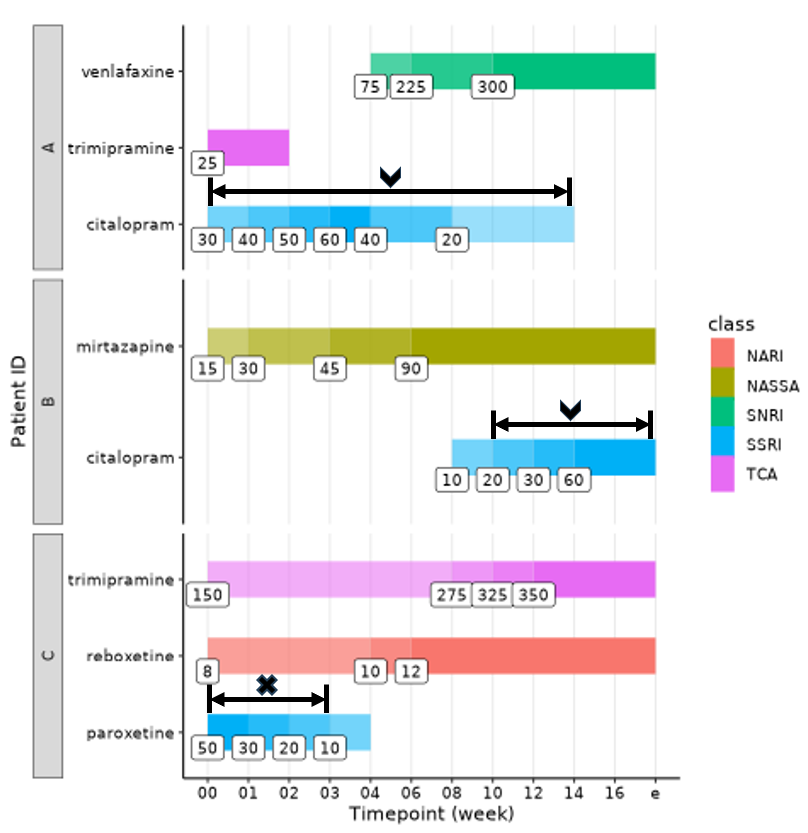

**Fig. S1:** **Timeline of antidepressant medication for three indicative patients in MARS.** Horizontal bars indicate the treatment duration for each antidepressant. Numbers within boxes represent the dosage (mg/day) administered at each timepoint. Colour shading corresponds to dosage intensity, with darker shades indicating higher doses.

Patient A received adequate dosages of citalopram from admission week through treatment week 14 and was therefore included as an SSRI user. Their percentage improvement was calculated based on HAMD-21 scores on the admission week and week 14. Patient B received adequate dosages of citalopram from treatment week 8 until discharge, so was included as an SSRI user. Their percentage improvement was calculated based on HAMD-21 scores on week 8 and prior to discharge. Patient C received adequate dosage of paroxetine for only 3 weeks, so was not included as an SSRI user.

NARI, Noradrenaline Reuptake Inhibitors; NaSSA, Noradrenaline and Specific Serotonergic Antidepressant; SNRI, Serotonin Noradrenaline Reuptake Inhibitors; SSRI, Selective Serotonin Reuptake Inhibitors; TCA, Tricyclic Antidepressants.

DAST

The Depression and Sequence of Treatment (DAST) study enrolled unrelated European patients diagnosed with MDD or bipolar disorder experiencing a major depressive episode. Participants were admitted for inpatient treatment at the University of Muenster, Germany, initiated at 2004 [4,5]. Inclusion criteria required a HAMD-21 admission score > 10 and a treatment duration of at least 6 weeks. The diagnosis of MDD was ascertained using the Structured Clinical Interview for Axis I Disorders based on DSM-IV criteria. Patients with schizoaffective disorders, comorbid substance abuse disorders, mental retardation, neurological or neurodegenerative disorders that could impair psychiatric evaluation were not included.

The study was approved by the University of Muenster ethics committee, and all participants provided written informed consent.

Patients were treated in a naturalistic setting with a variety of antidepressant medications, including citalopram/escitalopram, venlafaxine, mirtazapine, combination of these, and others (TCAs, MAOIs). Some patients also received co-medication with antipsychotics and mood stabilisers. Antidepressants have been regularly switched during week 1, and treatment response was assessed weekly based on intra-individual changes from the end of week 1 onward.

A total of 621 participants were recruited in the study, of which 586 were genotyped. Among them, 121 participants were treated with SSRIs and had information on non-remission and percentage improvement.

GENDEP

The Genome-based Therapeutic Drugs for Depression (GENDEP) project was a 12-week partially randomised open-label multi-centre pharmacogenetic study with two active treatment arms [6]. European patients with unipolar depression of at least moderate severity, diagnosed according to ICD-10 or DSM-IV criteria, were recruited across nine European centres. The diagnosis of MDD was ascertained using the semi-structured Schedules for Clinical Assessment in Neuropsychiatry (SCAN) interview. Patients with a personal or family history of schizophrenia or bipolar disorder and current substance dependence were not included.

The study was approved by the ethics boards of all participating centres, and all participants provided written informed consent.

Patients without contraindications were randomly assigned to flexible dosage of nortriptyline or escitalopram for 12 weeks, while those with contraindications for one drug were offered the other. Depression severity was assessed weekly by three rating scales: the clinician-rated MADRS as the primary one, and HAMD-17 and BDI for baseline assessment.

A total of 868 participants were recruited in the study, of which 796 were genotyped. Among them, 430 participants were treated with SSRIs and had information on non-remission and percentage improvement.

GenPod

The GENetic and clinical Predictors Of treatment response in Depression (GenPod) trial was a 12-week multi-centre randomised controlled trial with two treatment arms, initiated across three UK centres in 2008 [7]. Inclusion criteria required a diagnosis of an ICD-10 depressive episode (F32) using the Clinical Interview Schedule-Revised (CIS-R) and a BDI score ≥ 15 based on DSM-IV criteria. Patients who had taken antidepressant medication within two weeks prior to baseline assessment, had medical contraindications, or had a history of psychosis, bipolar disorder, or major substance or alcohol abuse were excluded.

Full ethical approval was obtained from the South West Ethical Board, along with research governance approval from the corresponding Primary Care Trusts. All participants provided written informed consent.

Patients were randomly allocated to receive either reboxetine or citalopram. Clinical outcomes were recorded at weeks 6 and 12 following randomisation. While the 12-week follow-up was carried out as an indicator of a persistent response to treatment, the primary outcome was assessed at week 6, when the maximum treatment response was expected.

A total of 601 participants were recruited, of which 477 were genotyped. Among them, 236 participants received citalopram and had information on non-remission and percentage improvement.

GODS

The Geneva Outpatient Depression Study (GODS) was a partially randomised trial [8]. Outpatients of various ethnicities diagnosed with moderate to severe depression according to DSM-IV-R and ICD-10 criteria, as established with the Mini International Neuropsychiatric Interview (MINI), were recruited between 1999 and 2002. Inclusion criteria required a baseline MADRS score ≥ 25. Patients with schizophrenia or schizoaffective disorder, current psychotic symptoms, dependence on alcohol or other substances within the preceding year, prior failure of treatment with one of the antidepressants used in the study, fluoxetine, monoamine oxidase inhibitors, mood stabilizers or antipsychotics were excluded.

The study protocol was approved by the ethics committee of the Geneva University Department of Psychiatry, and all participants provided written informed consent.

Depression severity was assessed at baseline and then bi-weekly using the MADRS. The GODS design allowed up to seven sequential treatment steps, based on the MADRS score at each visit. Patients remained on the same treatment step if they achieved a reduction of 25% (week 2), 40% (week 4), or 50% (week 6 and every four weeks thereafter) compared with baseline, otherwise treatment was stepped up. Patients were discharged only if complete remission was achieved. Step 1 began with 20 mg of paroxetine for all patients; step 2 increased to 30 mg of paroxetine. Step 3 introduced three randomised arms: 40mg of paroxetine (extended to 40 mg of paroxetine plus lithium at step 4); 150 mg of venlafaxine (extended to 300 mg at step 4); or 30mg of paroxetine plus lithium (extended to 40 mg plus lithium at step 4). From step 5 onward, randomisation ceased: non-responders received 150 mg of clomipramine, followed by clomipramine plus lithium at step 6, and clomipramine plus lithium plus triiodothyronine at step 7.

A total of 131 participants were recruited in the initial study, whereas 73 of which were genotyped and had information on non-remission or percentage improvement.

Data from GSK and Pfizer were shared by the Novel methods leading to new medications in depression and schizophrenia (NEWMEDS) Consortium.

NEWMEDS (GSK)

The Glaxo Smith Kline (GSK) samples came from two identically designed, randomised, double-blind, placebo-controlled trials comparing the antidepressant efficacy and effects on sexual functioning of bupropion XL and escitalopram in outpatients with moderate to severe depression, initiated in 2003 in the United States [9,10]. Diagnosis was based on DSM-IV criteria and assessed with a full psychiatric interview aided by MINI. Inclusion criteria required a current diagnosis of MDD lasting between 12 weeks to 2 years, a baseline HAMD-17 score ≥ 19, and having failed to respond to two adequate antidepressant trials within the previous 2 years. Patients were excluded if they had a history of current diagnosis of anorexia nervosa, bulimia, seizure disorder, or brain injury; a diagnosis of panic disorder, obsessive-compulsive disorder, posttraumatic stress disorder, or acute stress disorder within 12 months before study entry; a diagnosis of bipolar I or II disorder, schizophrenia, or other psychotic disorders; or a suicide attempt within 6 months before screening.

The protocol for the two trials was approved by international review boards, and all patients provided written informed consent.

The trials consisted of a 1-week screening phase followed by an 8-week treatment phase. On day 0 of the treatment phase, patients were randomly assigned to receive bupropion, escitalopram, or placebo. Clinical visits occurred at screening, randomisation (day 0), and treatment weeks 1,2,3,4,6, and 8.

A total of 210 patients were enrolled in the escitalopram treatment arms, of which 137 were of European ancestry and genotyped.

NEWMEDS (Pfizer)

The Pfizer cohort included patients from eight clinical trials of MDD. Study designs varied but were primarily double-blind, placebo-controlled trials lasting 6 to 8 weeks with sertraline, fluoxetine, or paroxetine [10]. Diagnosis and inclusion criteria required a screening HAMD-17 score ≥ 22. Patients with a DSM-IV diagnosis of psychotic features, bipolar I or II, or major risk for suicide were excluded.

All study protocols received institutional review board approval, and all participants provided written informed consent.

A total of 345 participants were enrolled, of which 311 were genotyped.

GSRD

The European Group for the Study of Resistant Depression (GSRD) was a multicentre, multinational, cross-sectional study with retrospective assessment of treatment response [11]. The study was conducted at 10 academic sites across eight European countries (Austria, Belgium, France, Germany, Greece, Israel, Italy, and Switzerland). Inpatients and outpatients with a diagnosis of MDD based on DSM-IV criteria, confirmed by the MINI, who had received ≥ 1 antidepressant trial during their current MDD episode prior to study entry (≥ 4 weeks at an adequate dose) were enrolled. Patients were excluded if they had any current primary psychiatric disorder other than MDD, or any substance disorder (except nicotine and caffeine) within the previous six months, or any concurrent severe personality disorder.

The study was approved by the ethics committees of all recruiting sites, and all participants provided written informed consent.

For each participant, two MADRS scores were estimated: a score at the timepoint of the cross-sectional data collection process (‘current MADRS’) and a retrospective MADRS score measuring the symptom severity at the onset of the current MDD episode. MADRS total score change during the current MDD episode was calculated as a measurement for treatment response. Antidepressant treatment followed naturalistic best-clinical practice principles.

A total of 1410 participants were enrolled, of which 734 received SSRI treatment—402 with augmentation or combination therapy and 332 with monotherapy. Of these SSRI users, 573 were genotyped.

PGRN-AMPS

The Pharmacogenomic Research Network Antidepressant Medication Pharmacogenomic Study (PGRN-AMPS) was an open-label trial with two treatment arms, citalopram and escitalopram [12]. Both inpatients and outpatients with nonpsychotic MDD and a baseline HAMD-17 score ≥ 14 were recruited at Mayo Clinic in Rochester, Minnesota, USA. Diagnosis was confirmed using the Structured Clinical Interview for DSM-IV (SCID). Patients were excluded if they had medical contraindications to citalopram or escitalopram, prior non-response to an adequate course of either drug, schizophrenia, schizoaffective disorder, bipolar I disorder, active substance abuse, suicidal ideation, or pregnancy.

The Institutional Review Board of Mayo Clinic approved and monitored the protocol, and all participants provided written informed consent.

Following baseline evaluation, patients were treated with 20 mg/day of citalopram or 10 mg/day escitalopram for eight weeks. Patients were evaluated at face-to-face follow-up at weeks 4 and 8 primarily using QIDS-C16. If the QIDS-C16 score was ≤ 5, the dose was maintained; if between 6 and 8, clinicians would evaluate the subject and either maintain or increase the dose; and if ≥ 9, the dose was increased unless contraindicated by intolerable side effects or patients’ refusal. The scheduled dose increase at the four-week visit was to 40 mg/day for citalopram or 20 mg/day for escitalopram.

A total of 529 participants were enrolled and genotyped.

Mayo

The Mayo cohort was a continuation and replication of the PGRN-AMPS cohort [13]. Patients were recruited using the same protocol and inclusion criteria as described above for PGRN-AMPS, but genotyping was carried out separately, and the primary depression rating scale was HAMD-17.

**Mixed ancestry**

STAR*D

The Sequenced Treatment Alternatives to Relieve Depression (STAR*D) study was a multisite, prospective, sequentially randomised clinical trial of outpatients with nonpsychotic MDD conducted across 18 primary care and 23 psychiatric clinical sites in the United States [14–16]. Inclusion criteria required a diagnosis of nonpsychotic MDD confirmed using a DSM-IV checklist and a baseline HAMD-17 score ≥ 14. Patients were excluded if they had a prior inadequate response to a robust trial to any of the protocol treatments, a history of clear-cut intolerance to a protocol treatment during the current episode, a primary diagnosis of schizophrenia, schizoaffective disorder, anorexia, bulimia, or obsessive-compulsive disorder, substance dependence, or were pregnant or breast-feeding.

The University of Texas Southwestern Medical Centre at Dallas and the institutional review boards at each clinical site and regional centre and the Data Coordinating Centre and the Data Safety and Monitoring Board of the National Institute of Mental Health (NIMH) approved and monitored the protocol, and all participants provided written informed consent.

At level 1, all patients received flexible doses of citalopram for up to 14 weeks. Those without sufficient symptomatic benefit were randomly assigned to level 2 treatments, which entailed four switch options (sertraline, bupropion, buspirone, or cognitive psychotherapy) and three citalopram augmentations (bupropion, buspirone, or cognitive psychotherapy). Patients who received cognitive therapy without sufficient improvement were randomly assigned to level 2A switch options (venlafaxine or bupropion). Level 2 or 2A patients without sufficient improvement after at least two medications were randomly assigned to level 3 treatments, which included two switch options (mirtazapine or nortriptyline) or two augment options (lithium or L-triiodothyronine). Level 4 involved random assignment to two switch options: tranylcypromine or mirtazapine plus venlafaxine.

The primary outcome of the original study was measured by HAMD-17 via telephone-based structured interviews at study entry and at the end of each level. Secondary outcomes were based on the QIDS-SR collected at baseline and at each treatment visit at weeks 2,4,6,9,12, and optionally 14, where analysis primarily relied on due to its data richness for most participants.

A total of 4790 participants were screened, of which 2876 were eligible for analysis, and 1948 were genotyped. To maximize power for association analysis, we focused on Level 1 data only, where all participants received citalopram.

**East Asian ancestry**

SEOUL_DEP

The SEOUL_DEP cohort recruited patients with MDD of unrelated Korean ancestry from the clinical trials program of the Samsung Medical Centre Geropsychiatry and Affective Disorder Clinics, conducted in a naturalistic clinical setting [17]. Inclusion criteria required a diagnosis of a current unipolar major depressive episode according to DSM-IV, established through an initial clinical interview, followed by a structured research assessment using the Samsung Psychiatric Evaluation Schedule, which includes the Structured Clinical Interview for DSM-IV. The diagnosis was finalised by a board-certified psychiatrist after reviewing clinical observations, medical records, history and the Structured Clinical Interview for DSM-IV interview. A baseline HAMD-17 score ≥ 15 was also required. Exclusion criteria were pregnancy, significant medical conditions, abnormal laboratory baseline values, suicide attempt, history of alcohol or drug dependence, seizure, neurological illness including significant cognitive impairment, or any concomitant DSM-IV Axis I psychiatric disorder.

The ethics review board of Samsung Medical Centre approved the study, and all participants provided written informed consent.

Patients received monotherapy with escitalopram, sertraline, fluoxetine, paroxetine, or mirtazapine for 6 weeks. The clinician’s antidepressant choice was naturalistic, considering anticipated adverse effects. Dose titration was completed within 2 weeks. HAMD-17 assessments were administered by a single trained rater every 2 weeks.

Only participants treated with one of the four SSRIs were included in the current analysis, totalling 555 participants.

The Japan, Miaoli, and Taipei cohorts were included in the International SSRI Pharmacogenomics Consortium (ISPC) GWAS of antidepressant response and has been previously described [18]. The primary depression rating scale was HAMD-21 for all these three cohorts, but only 17 items in align with the HAMD-17 were summed into the total score for analysis of antidepressant treatment outcome.

Japan

The Japan cohort was an open-label, randomised trial with two treatment arms, paroxetine and fluvoxamine, conducted at the Department of Neuropsychiatry, Kansai Medical University, initiated in 2002 [19]. Japanese patients meeting DSM-IV criteria for recurrent MDD (excluding bipolar disorder) and free of psychotic drugs except benzodiazepines for more than 10 days were included. The diagnoses were made by two independent senior psychiatrists and confirmed by a third psychiatrist who was blinded to the previous evaluations, using Structured Clinical Interview for DSM-IV Axis I Disorders. Patients were excluded if they had clinically significant unstable medical illness, pregnancy, a primary psychiatric diagnosis other than MDD, or had received electroconvulsive therapy within the previous 6 months.

The ethics committee of Kansai Medical University and Osaka University approved the study, and all participants provided written informed consent.

Treatment response was evaluated at baseline and after 2, 4, and 6 weeks of antidepressant treatment.

A total of 121 participants were enrolled and genotyped.

Miaoli

The Miaoli cohort recruited outpatients from five hospitals in northern Taiwan to evaluate the efficacy and tolerability of escitalopram versus paroxetine [20,21]. Inclusion criteria required an MDD diagnosis according to DSM-IV criteria, confirmed using the Structured Clinical Interview for DSM-IV Axis-I disorders, conducted by board-certified psychiatrists and trained research nurses, and a baseline HAMD-21 score ≥ 14. Patients were excluded if they had a primary or comorbid diagnosis of schizophrenia, schizoaffective disorder, bipolar disorder, alcohol or substance dependence, dementia, or other significant medical conditions, or if they had previously been treated with paroxetine or escitalopram.

The study was approved by the institutional review boards of the National Health Research Institutes and all participating clinics, and all participants provided written informed consent.

Before entering the study, patients completed a 7-day washout period for any prior antidepressant treatment (12 days for fluoxetine). Patients were administered either escitalopram or paroxetine according to the judgment of study clinicians. In the escitalopram arm, patients received a daily fixed dose of 10 mg for the first 4 weeks, followed by flexible dosing of 10–30 mg/day based on clinical response during an 8-week treatment period. In the paroxetine arm, patients received a daily fixed dose of 20 mg paroxetine for the first 4 weeks, and then a second 4-week period of flexible dosing (20–40 mg/day). No other psychotropic drugs were allowed during this period, except for 10 mg of zolpidem per night to treat insomnia (as needed, but for no more than four nights per week) and 1–2 mg of lorazepam per day to treat anxiety symptoms as needed. Study participants were assessed using the HRSD-21 and the Hamilton Rating Scale for Anxiety, at weeks 0, 1, 2, 4, 6, and 8 of continuous treatment.

A total of 245 participants were enrolled and genotyped.

Taipei

The Taipei cohort recruited outpatients of Chinese ethnicity with moderate-to-severe depression from a psychiatric clinic [22–25]. Inclusion criteria required a diagnosis of MDD according to DSM-IV guidelines, confirmed by a Structured Clinical Interview, a baseline HAMD-21 score ≥ 18, and the presence of depressive symptoms for at least 2 weeks without antidepressant treatment (patients were either new cases or had discontinued antidepressants for more than 2 weeks). Patients were excluded if they had additional DSM-IV Axis 1 diagnoses (including substance use, generalised anxiety disorder, panic disorder, or obsessive-compulsive disorder), personality disorder, pregnancy, a history of attempted suicide, or major medical and/or neurological disorders.

The Institution Review Boards in Veteran General Hospital-Taipei and E-DA Hospital approved the study, and all participants provided written informed consent.

Daily doses of fluoxetine or citalopram were administered, starting at 20 mg/day and could be increased to 40 mg/day based on clinical response. No other psychotropic medications were permitted; however, anxiolytics were allowed for insomnia. Treatment efficacy was evaluated at weeks 4 and 8.

A total of 230 psychiatric outpatients were enrolled, of which 177 participants had information on non-remission and percentage improvement, and 174 were genotyped.

### 2 Measures of antidepressant response

Specifically, non-remission was defined using these thresholds for each rating scale: Montgomery Åsberg Depression Rating Scale (MADRS) > 10, Quick Inventory of Depressive Symptomatology (QIDCS) > 5, 17-item Hamilton Depression Rating Scale (HAMD-17) > 7, 21-item Hamilton Depression Rating Scale (HAMD-21) > 7, and Beck Depression Inventory (BDI) > 9 [26–29].

Severe depression was defined using the following thresholds: BDI ≥ 29, HAMD-17/HAMD-21 ≥ 24; MADRS ≥ 35; QIDS-SR/QIDS-C ≥ 16 [30,31].

### 3 Descriptive statistics across cohorts

Since only GWAS summary statistics from the SEOUL_DEP cohort were shared with the PGC MDD Antidepressant Response Working Group, this cohort is not included in the following figures.

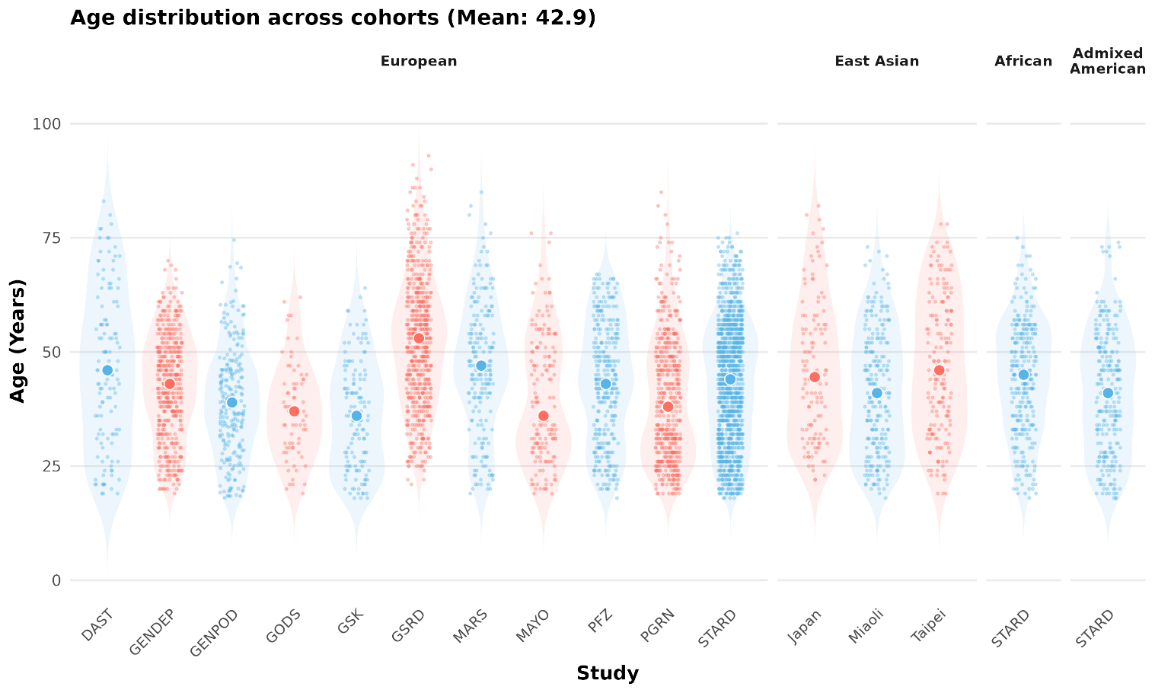

Figure S2 Age of study participants at recruitment, across cohorts

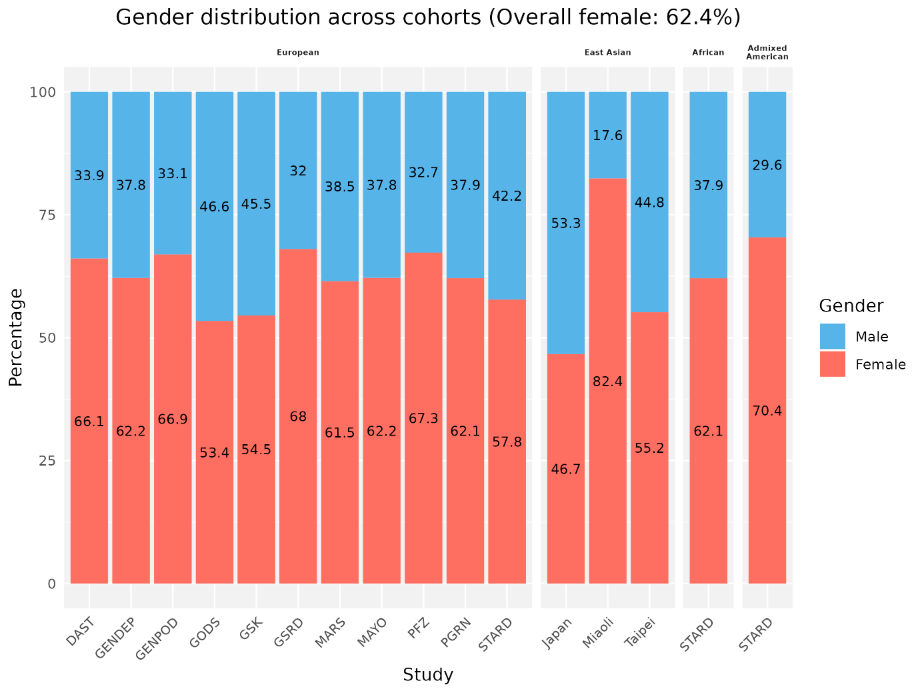

Figure S3 Proportion of study participants by gender across cohorts

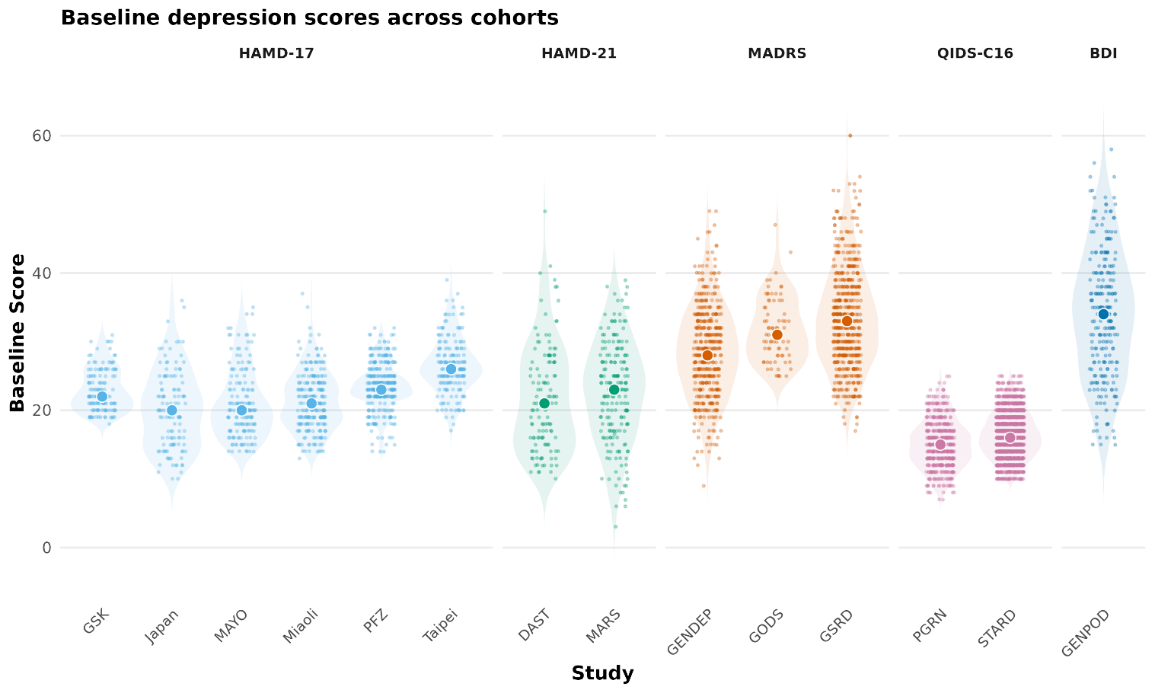

Figure S4 Baseline depression score across cohorts, by depression rating scale

### 4 Quality control and imputation using RICOPILI

Individual-level genotype data from all 14 cohorts were processed using the RICOPILI pipeline following PGC criteria, including standardised quality control (QC), principal component analysis (PCA), and imputation [32].

4.1 Technical Quality Control (QC) of the 14 cohorts

The default parameters of QC for retaining SNPs and subjects were: SNP call rate ≥ 0.95 (before sample removal), subject call rate ≥ 0.98, SNP call rate ≥ 0.98, deviation of autosomal heterozygosity from the mean (|Fhet|≤0.2), difference in SNP missingness between remitters and non-remitters ≤ 0.02, sample with discordant sex were excluded based on X chromosome inbreeding coefficient (F) (pedigree males with Fhet < 0.5 and females with Fhet > 0.5), SNP with valid association p-value, SNP minor allele frequency (MAF) ≥ 0.01, deviation from Hardy-Weinberg equilibrium (HWE, p > 1e-06 for remitters, p > 1e-10 for non-remitters).

4.2 Genomic Quality Control: Principal Component Analysis (PCA) and Relatedness Checking

Across all 14 cohorts, we performed PCA using autosomal SNPs with high imputation quality (INFO > 0.8), low missingness (<1%), and MAF > 0.05. SNPs were pruned to approximate linkage equilibrium using two iterations of LD pruning (r² < 0.2, 200-SNP windows), and known long-range LD regions (e.g., MHC and the chromosome 8 inversion) were excluded. A total of 36,957 SNPs in European-ancestry cohorts, 61,634 SNPs in East Asian-ancestry cohorts, 100,016 SNPs in African-ancestry cohort, and 63,303 SNPs in Admixed American-ancestry cohort, shared across all cohorts within each ancestry group and passing these filters, were used for robust relatedness testing using PLINK v1.9. Pairs of individuals with PIHAT > 0.2 were identified, and one individual from each pair was removed at random, with preference given to retaining cases over controls.

To control for false-positive associations arising from inflated test statistics, we evaluated the effectiveness of technical and genomic QC measures using the genomic inflation factor (λGC, median). QC parameters were iteratively tightened as needed until λGC was between 0.962 and 1.096 prior to inclusion of principal components (PCs), and between 0.954 and 1.026 after including PCs as covariates.

Twenty genetic principal components (PCs) were derived from the quality-controlled genotype data and evaluated using logistic regression. PCs 1-4 were included for all ancestries; additionally, PC5 was included for European ancestry, PCs 6 and 12 for East Asian ancestry, and PC20 for African ancestry to control for population stratification in downstream analyses. The contribution of individual PCs to genome-wide test statistics was further assessed using the genomic inflation factor (λGC).

The ancestry of samples was confirmed and assigned by comparison to the 1000 Genome reference using EIGENSTRAT [33]. Specifically, European ancestry of the 1000 Genome reference panel included Utah residents with Northern and Western European ancestry (CEU), Finnish in Finland (FIN), British from England and Scotland, UK (GBR), Iberian Populations in Spain (IBS), and Toscani in Italia (TSI); East Asian ancestry included Chinese Dai in Xishuangbanna, China (CDX), Han Chinese in Beijing, China (CHB), Han Chinese South, China (CHS), Japanese in Tokyo, Japan (JPT), and Kinh in Ho Chi Minh City, Vietnam (KHV); African ancestry included African Caribbean in Barbados (ACB), People with African Ancestry in Southwest USA (ASW), Esan in Nigeria (ESN), Gambian in Western Division, Mandinka (GWD), Luhya in Webuye, Kenya (LWK), Mende in Sierra Leone (MSL), and Yoruba in Ibadan, Nigeria (YRI); Admixed American ancestry included Colombians in Medellin, Colombia (CLM), People with Mexican Ancestry in Los Angeles, CA, USA (MXL), Peruvians in Lima, Peru (PEL), Puerto Ricans in Puerto Rico (PUR) [34].

4.3 Genotype imputation

Genotype imputation was performed using the reference-based phasing/state space reduction of the hidden Markov models (HMMs) approach implemented in Eagle2/ Minimac3 [35,36]. The reference panel consisted of 64,976 human haplotypes at 39,235,157 SNPs integrated from 20 studies of predominantly European ancestry from the Haplotype Reference Consortium (HRC) [37] . Chromosome X was imputed separately for males and females using the same HRC reference panel, with additional QC filters applied (MAF > 0.05, Hardy-Weinberg equilibrium p-value > 1e-6, missingness rate < 0.02). For males, invalid heterozygous haploid variants outside the Pseudoautosomal Regions (PAR) were set to missing. After imputation, we obtained the probability of each genotype for each SNP in each individual. We then derived best guess genotypes by selecting the genotype with the highest probability, applying a minimum threshold (probability P > 0.8), and performing stricter quality control (minor allele frequency (MAF) > 0.05, missingness rate < 0.01).

Within-ancestry PCA was then performed for each of the four ancestries by integrating best guess strict genotypes (MAF > 0.05, SNP missing rate < 0.02) from cohorts containing samples of the corresponding ancestry. For instance, PCA for African ancestry only included samples from STAR*D, while PCA for Asian ancestry included samples from Japan, Taipei, and Miaoli. This aimed to identify overlapping or related individuals across cohorts and to generate principal components for use as covariates in downstream association analyses. When related individuals were identified, cases (in this study, non-remitters) were given priority over controls (remitters) for retention.

### 5 QQ-plots for GWAS

|  | Remission | Percentage Improvement |
| --- | --- | --- |
| EUR | 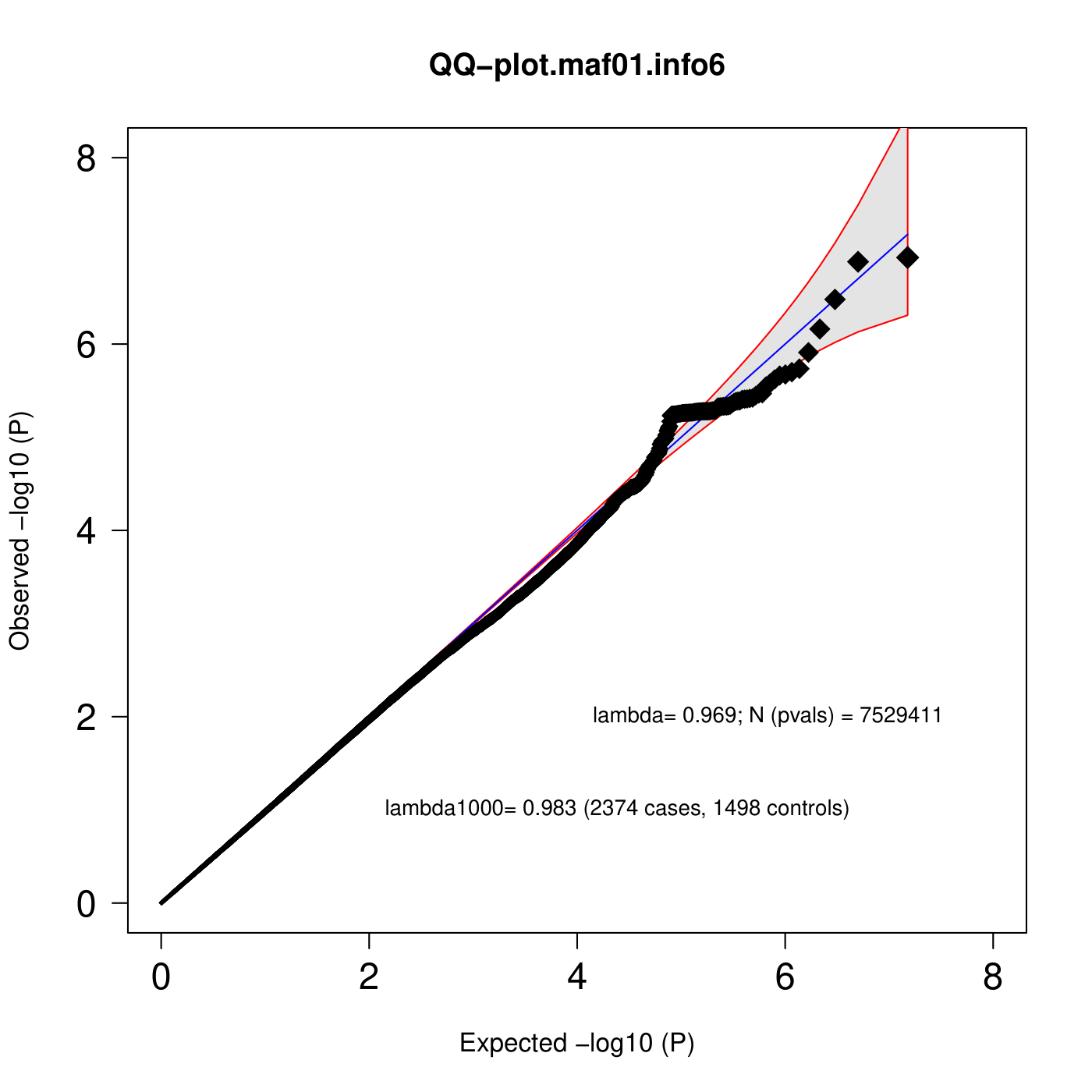 | 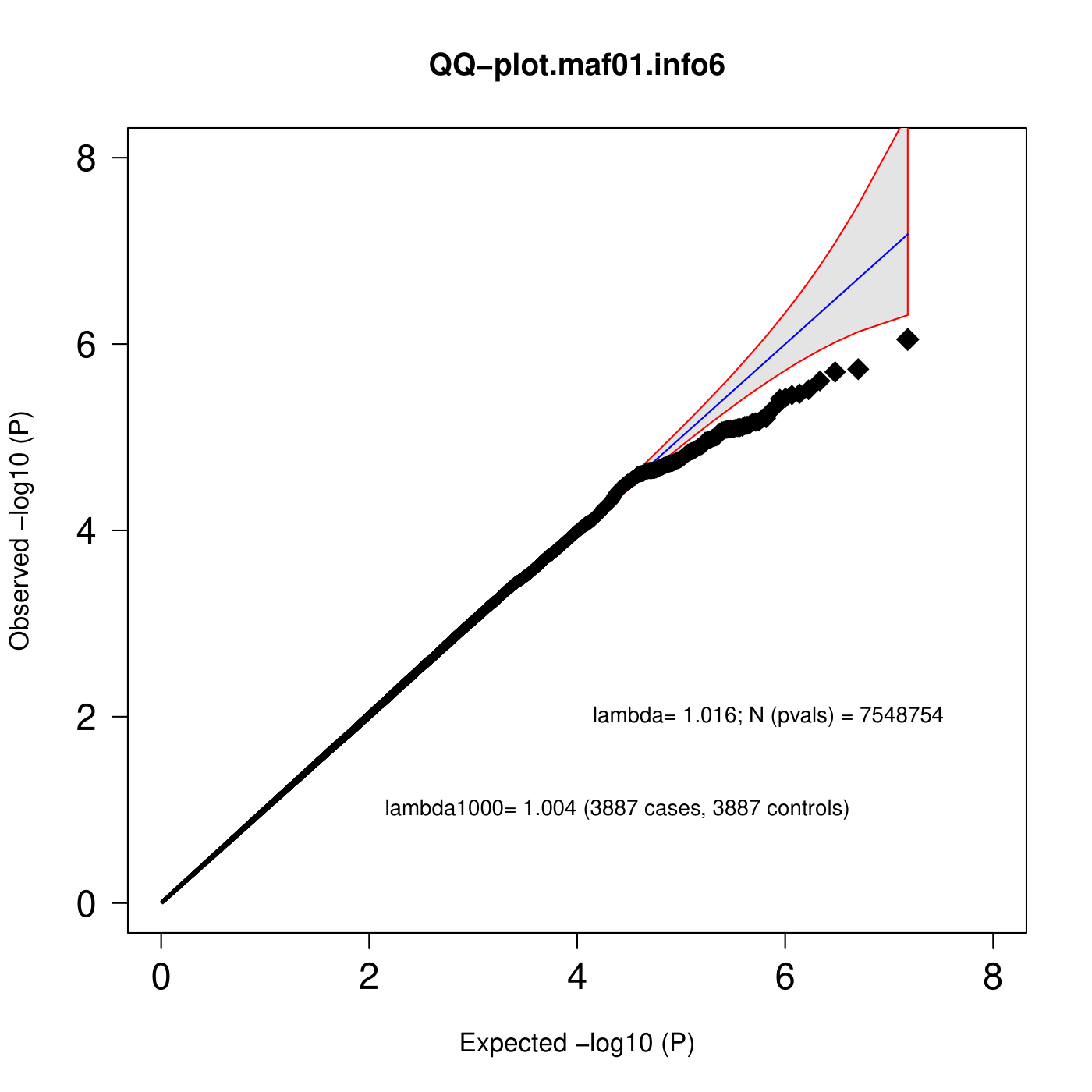 |
| EAS | 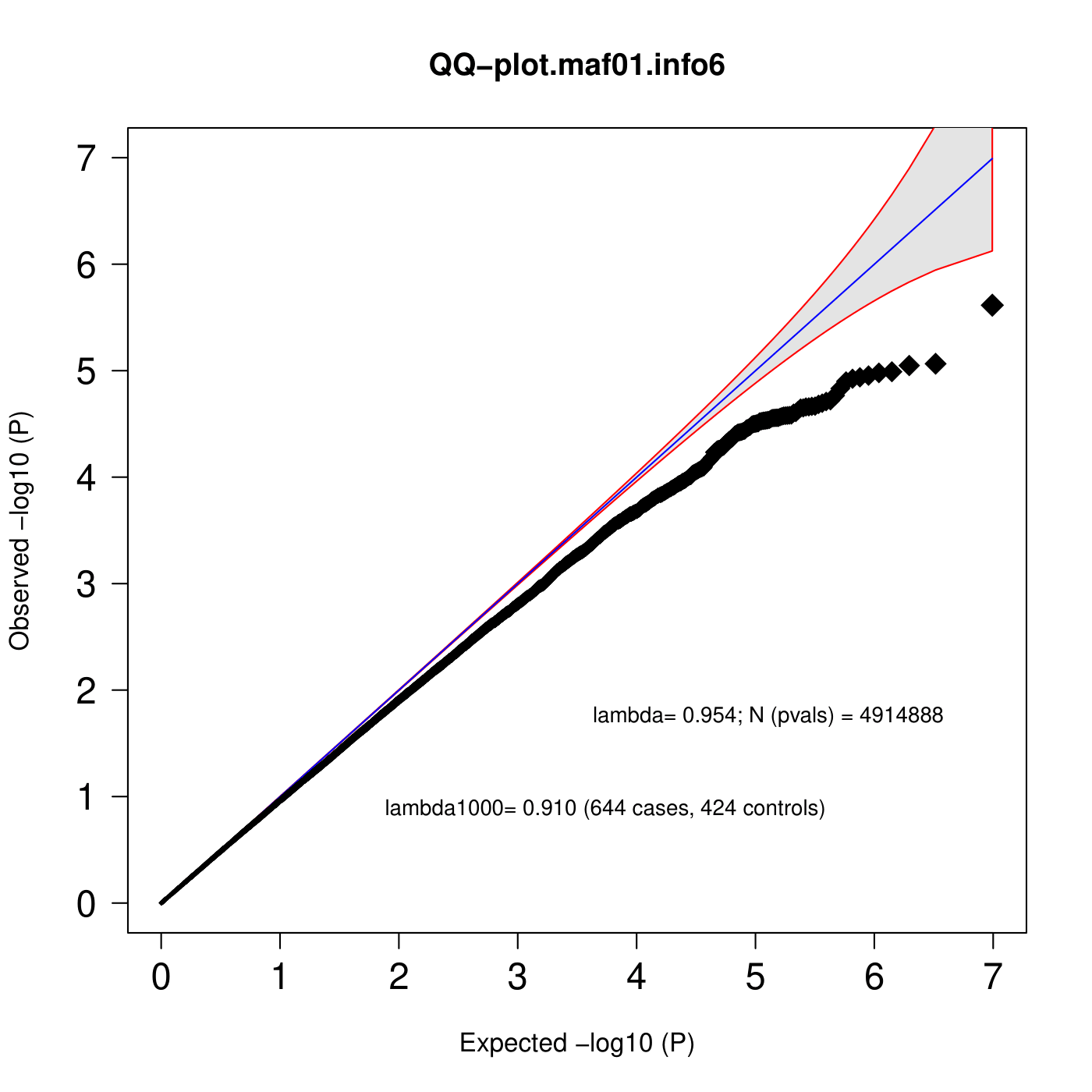 | 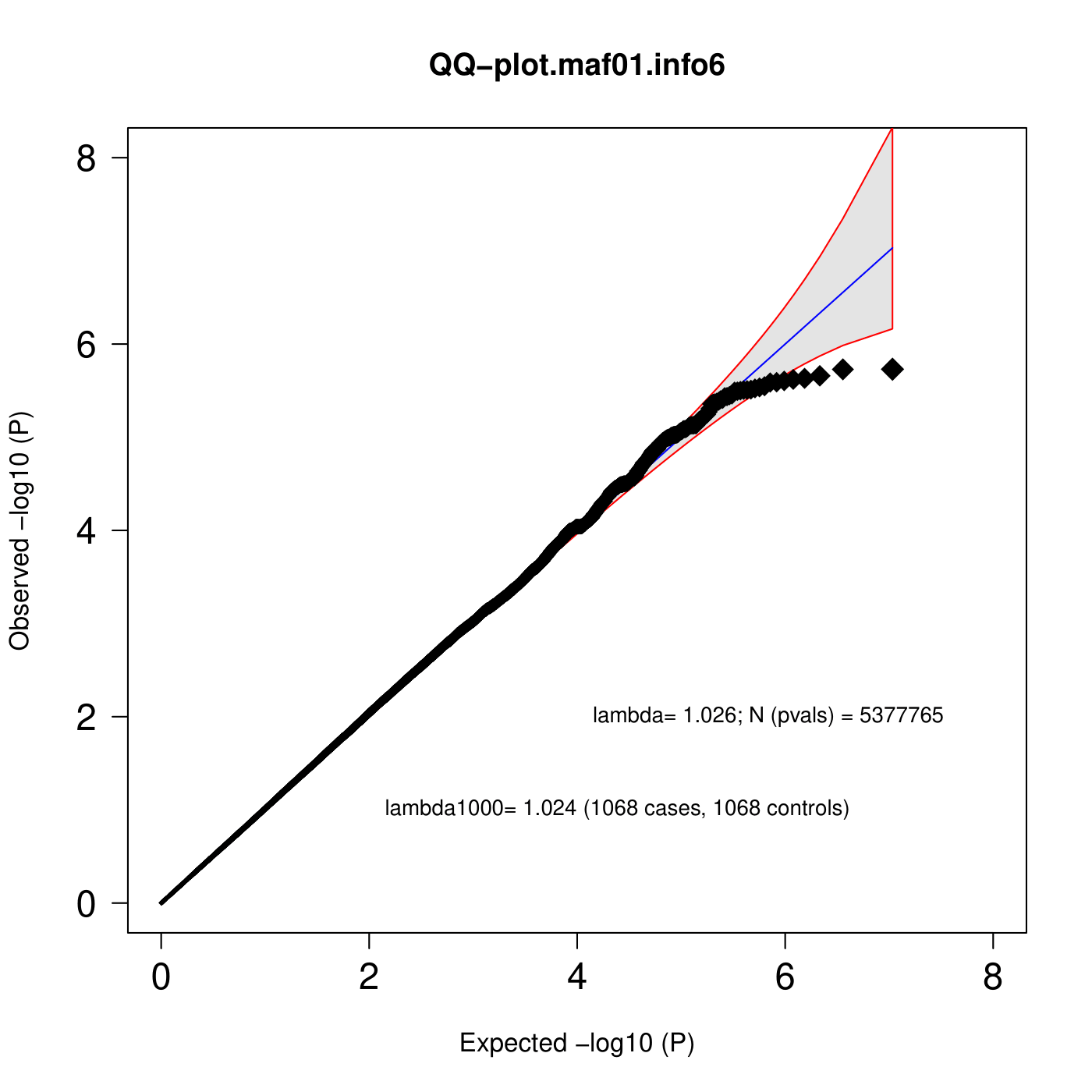 |
| AFR | 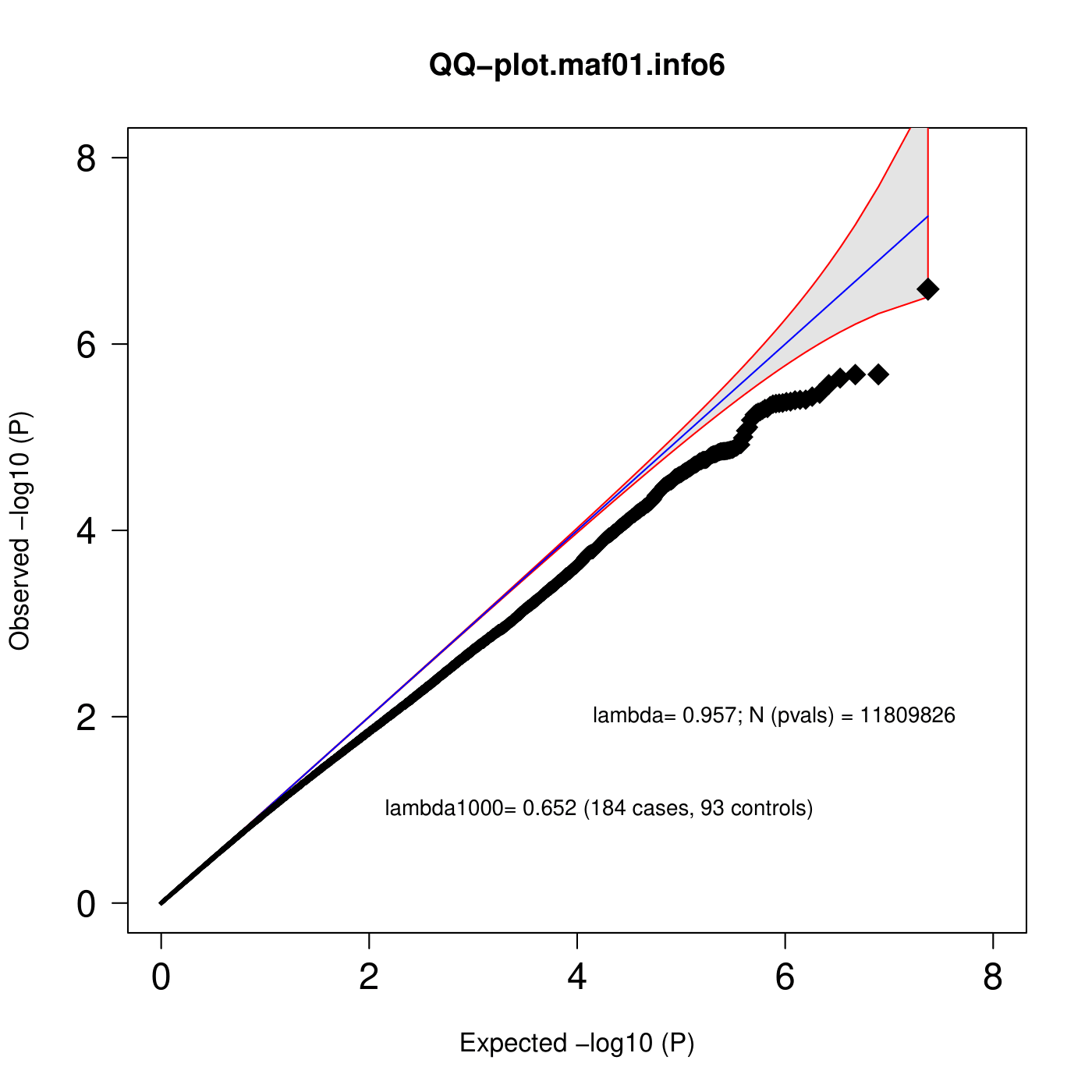 | 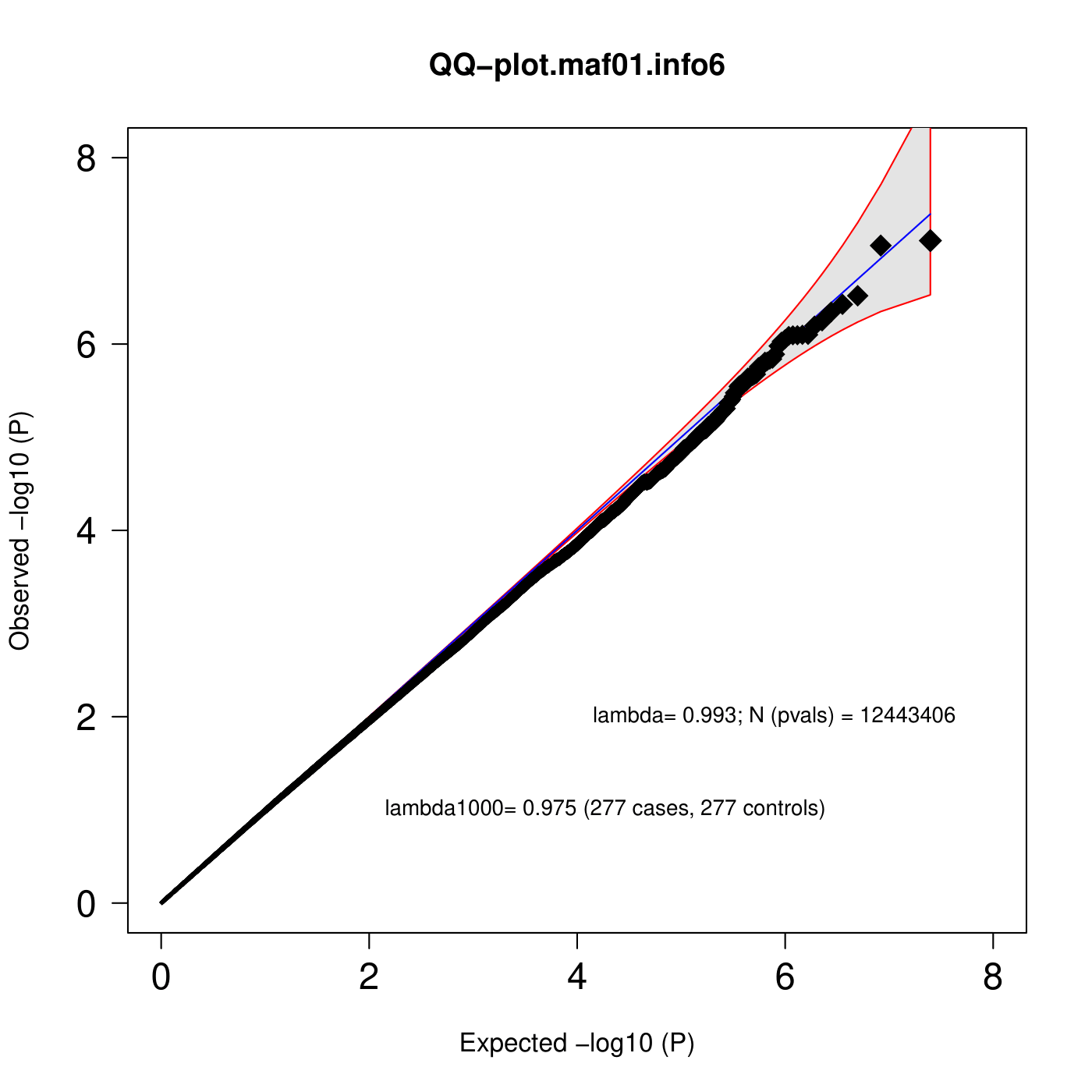 |
| AMR | 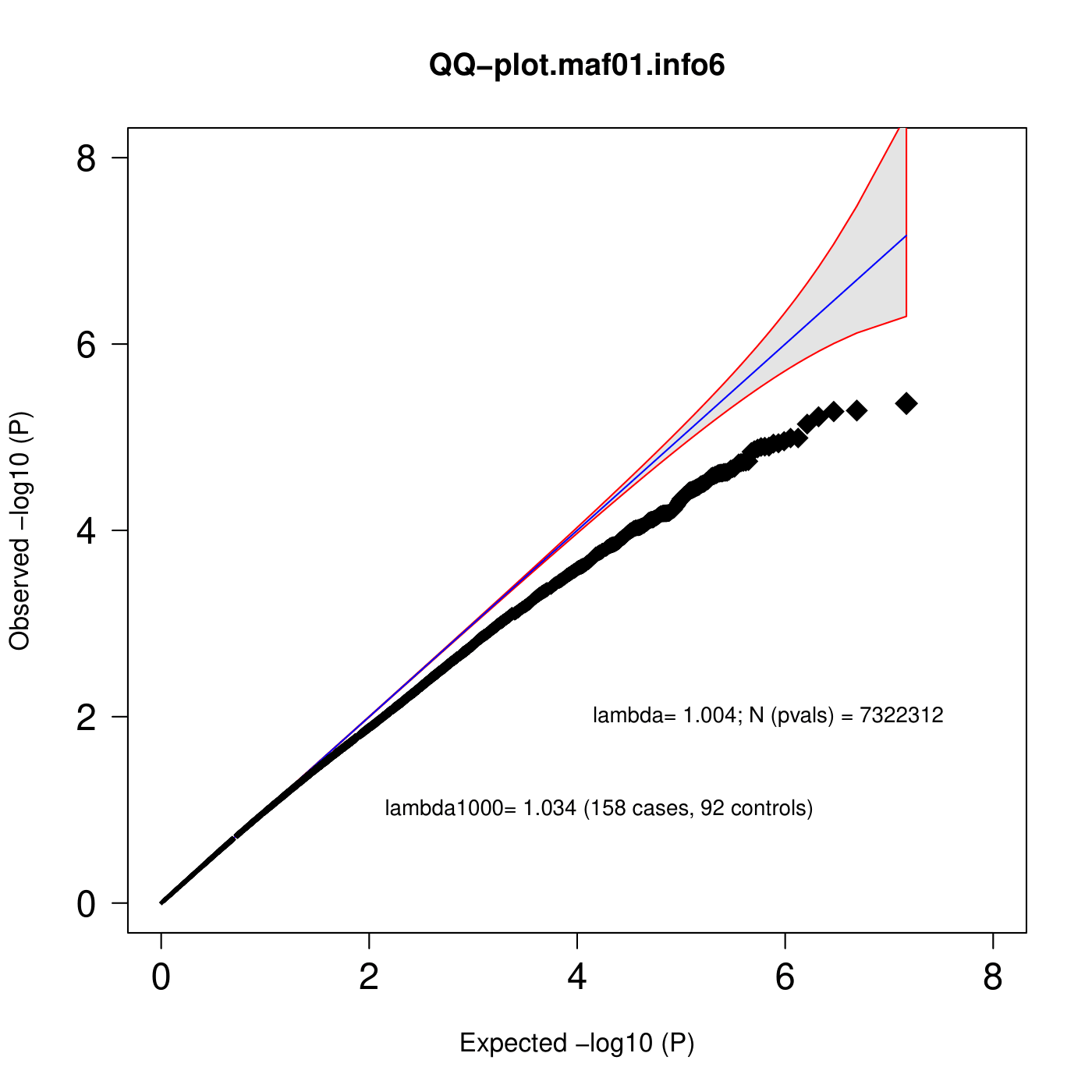 | 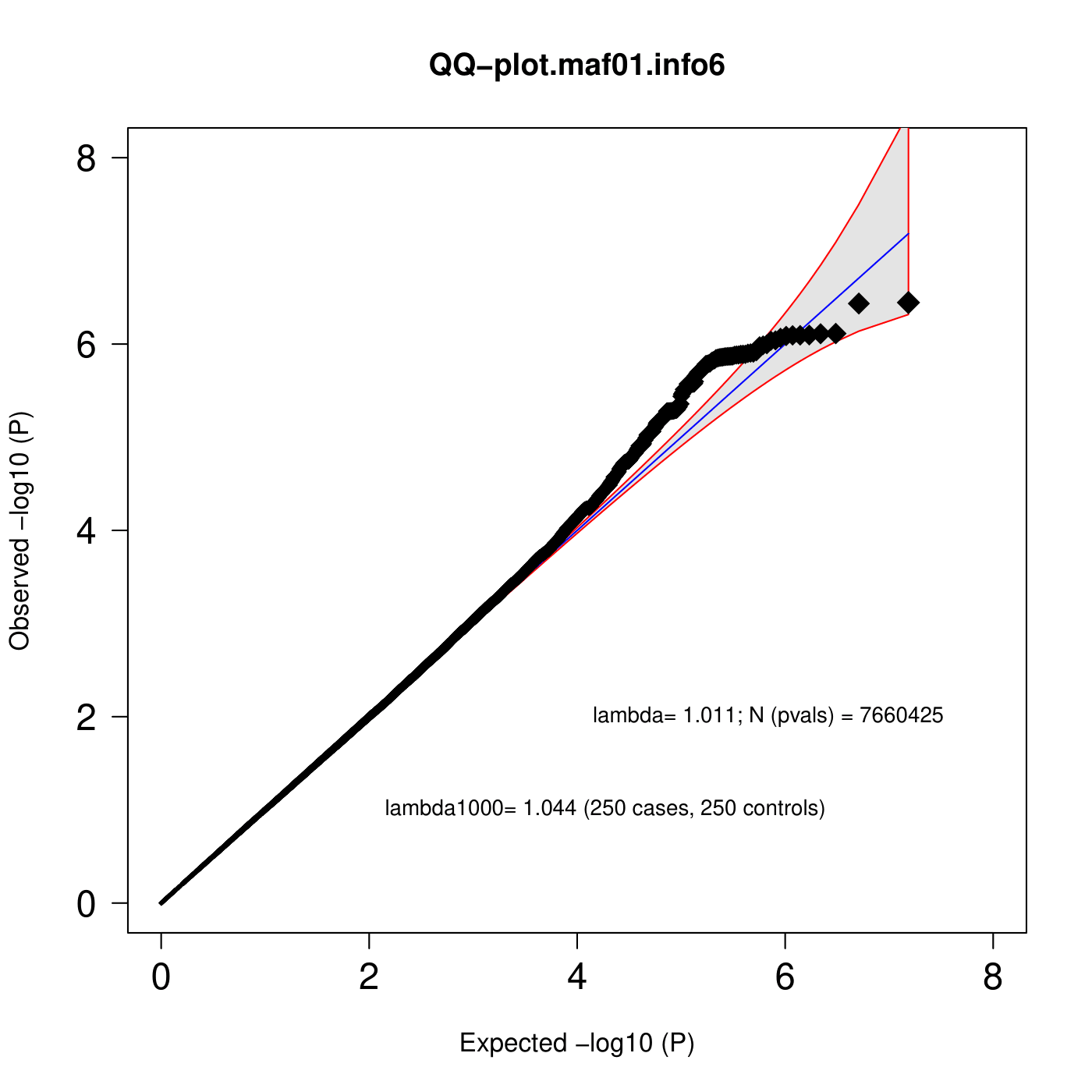 |
| Trans- | 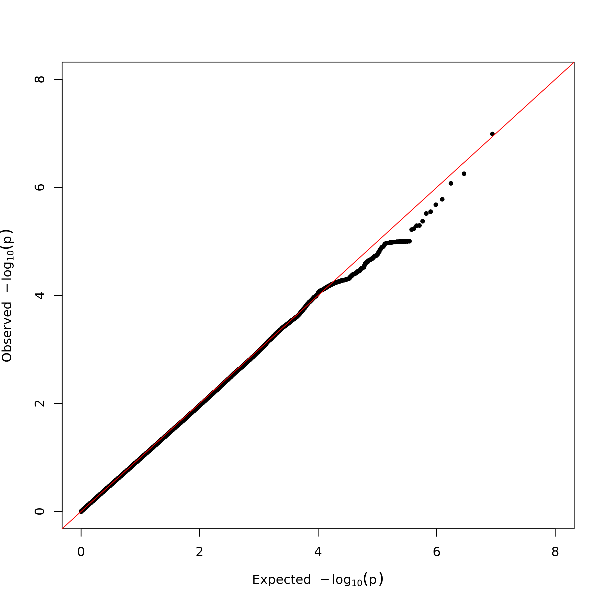 | 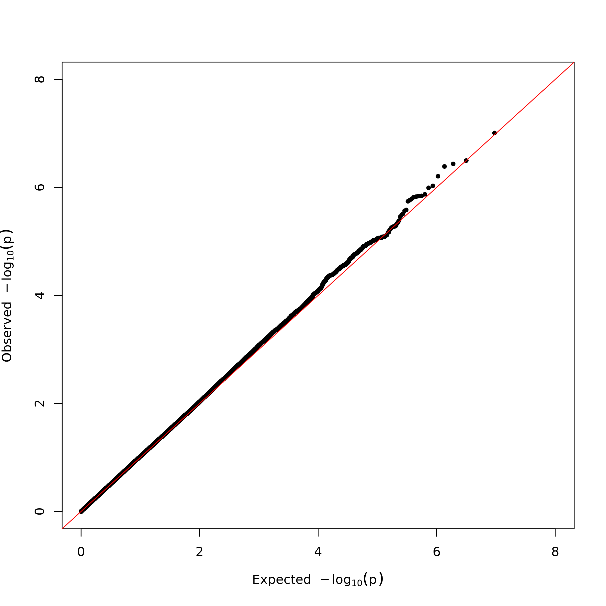 |
| Figure S5 QQ-plots for GWAS of non-remission (left) and percentage improvement (right) in each ancestry and trans-ancestry meta-regression | | |

### 6 SNP-based heritability estimation

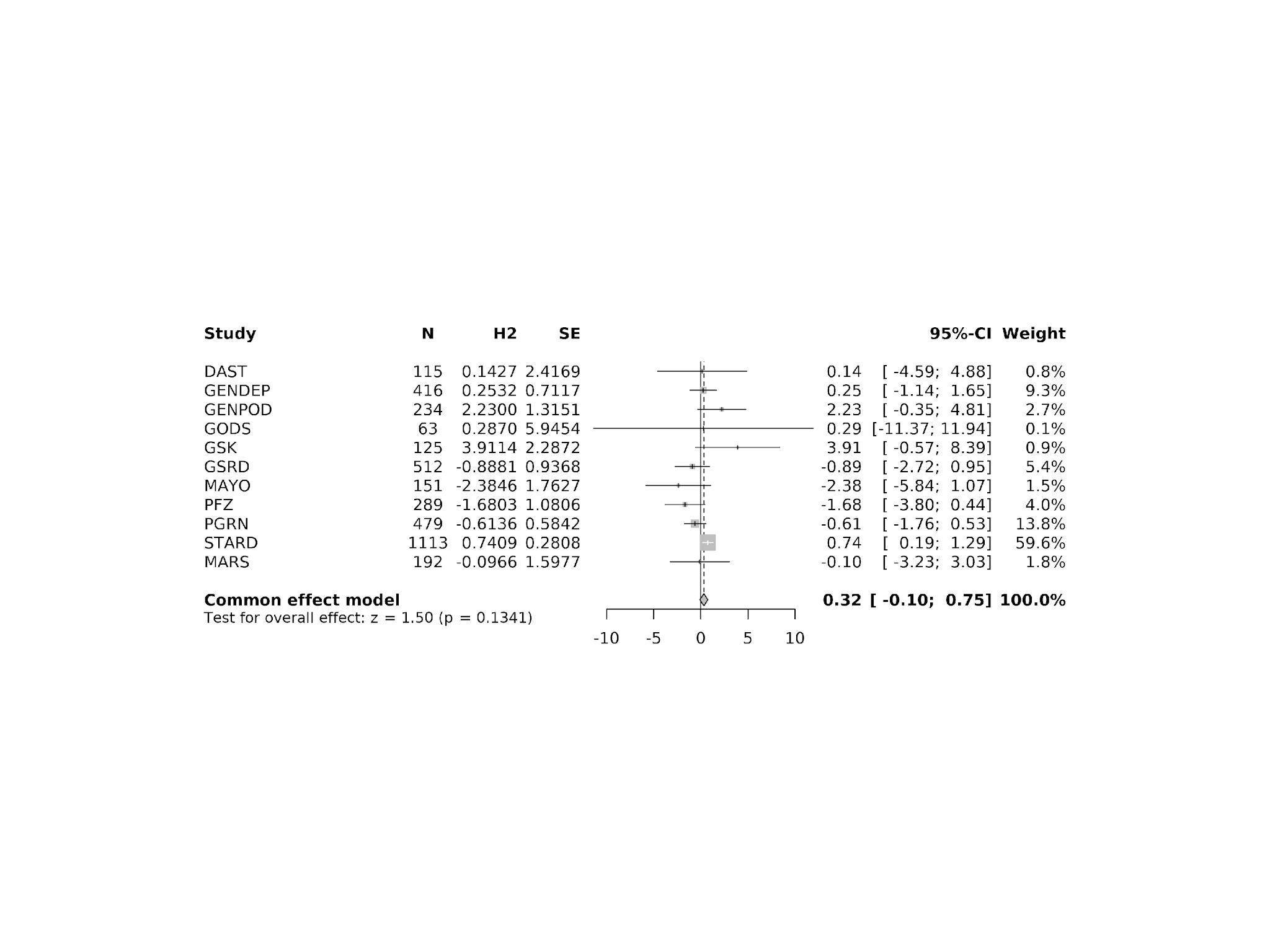

Figure S6 Forest plot showing meta-analysis of per cohort SNP-based heritability estimation for non-remission, in European ancestry studies, using GREML

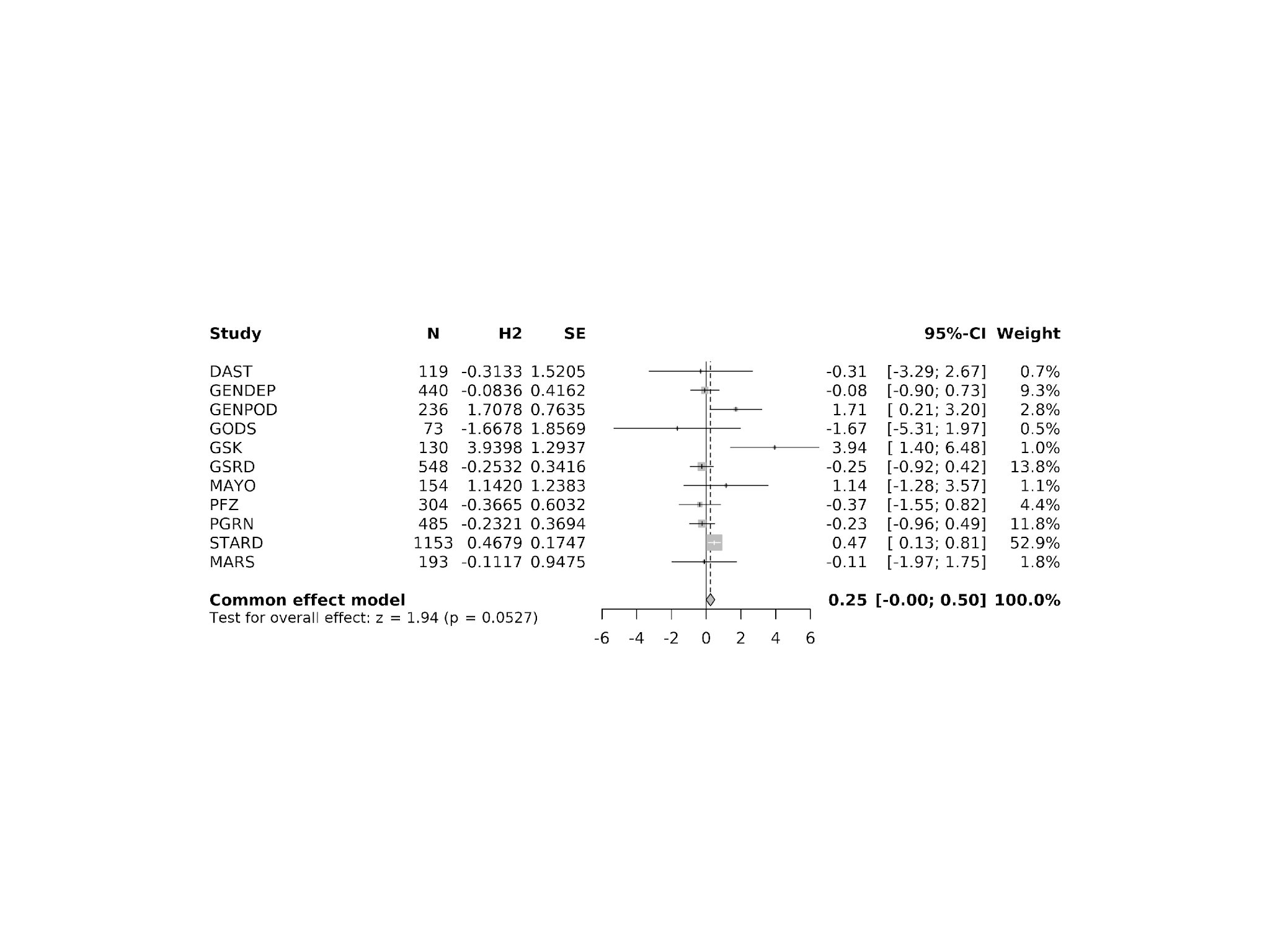

Figure S7 Forest plot showing meta-analysis of per cohort SNP-based heritability estimation for percentage improvement, in European ancestry studies, using GREML

### 7 Shared genetic architecture with 13 psychiatric traits

| Table S1 GWAS summary statistics of polygenic scoring prediction. N_eff_half, power equivalent to a case-control study of this size of cases and controls, calculated by 2×N_case×N_control/(N_case+ N_control) | | | | | |
| --- | --- | --- | --- | --- | --- |
| Phenotypes | PMID | Authors (Ref) | N_eff_half | Case_N | Control_N |
| Major Depressive Disorder | 39814019 | Adams et al., 2025 [38] | 1,004,459 | 688,808 | 4,364,225 |
| Anxiety Disorders | 39006447 | Strom et al., 2026 [39] | 195,264 | 122,341 | 729,881 |
| Bipolar Disorder | 39843750 | O'Connell et al., 2025 [40] | 535,720 | 158,036 | 2,796,499 |
| Schizophrenia | 35396580 | Trubetskoy et al., 2022 [41] | 79,740 | 76,755 | 243,649 |
| Post Traumatic Stress Disorder | 38637617 | Nievergelt et al., 2024 [42] | 345,433 | 150,793 | 1,130,197 |
| Obsessive-Compulsive Disorder | 38712091 | Strom et al., 2025 [43] | 25,981 | 23,493 | 1,114,613 |
| Anorexia nervosa | 31308545 | Watson et al., 2019 [44] | 23,160 | 16,992 | 55,525 |
| Treatment Resistant Depression | 33753889 | Fabbri et al., 2021 [45] | 3,725 | 2,146 | 14,097 |
| Attention Deficit/Hyperactivity Disorder | 36702997 | Demontis et al., 2023 [46] | 51,568 | 38,691 | 186,843 |
| Autism Spectrum Disorder | 30804558 | Grove et al., 2019 [47] | 22,183 | 18,381 | 27,969 |
| Electronic health record SSRI switching | 40510220 | Lo et al., 2024 [48] | 8,908 | 5,133 | 33,680 |
| SSRI non-response | 41261141 | Koch et al., 2025 [49] | 38,854 | 21,956 | 108,364 |
| Lithium response (for bipolar disorder) | 37433967 | Amare et al., 2023 [50] | 952 | 660 | 1,707 |

| Table S2 Regression results between polygenic scores (PGS) for 13 psychiatric traits and SSRI response | | | | | | | |
| --- | --- | --- | --- | --- | --- | --- | --- |
|  | Non-remission | | |  | Percentage Improvement | | |
|  | Odds ratio (OR) | 95% CI | p |  | β | 95% CI | p |
| Attention-deficit/hyperactivity disorder | 1.054 | [0.982, 1.130] | 0.144 |  | -0.009 | [-0.041, 0.024] | 0.591 |
| Anorexia nervosa | 1.007 | [0.943, 1.075] | 0.832 |  | 0.014 | [-0.017, 0.044] | 0.371 |
| Anxiety | 1.069 | [1.000, 1.143] | 0.051 |  | -0.023 | [-0.054, 0.008] | 0.146 |
| Autism spectrum disorder | 0.994 | [0.928, 1.066] | 0.874 |  | 0.011 | [-0.021, 0.043] | 0.505 |
| Bipolar disorder | 1.022 | [0.955, 1.093] | 0.535 |  | -0.017 | [-0.048, 0.015] | 0.302 |
| Major depressive disorder | 1.074 | [1.002, 1.151] | 0.042 |  | -0.035 | [-0.067, -0.003] | 0.032 |
| Obsessive-compulsive disorder | 0.993 | [0.929, 1.061] | 0.829 |  | 0.007 | [-0.024, 0.038] | 0.662 |
| Post-traumatic stress disorder | 1.085 | [1.013, 1.162] | 0.021 |  | -0.032 | [-0.064, 0.000] | 0.051 |
| Schizophrenia | 1.088 | [1.013, 1.168] | 0.020 |  | -0.022 | [-0.055, 0.011] | 0.195 |
| SSRI switching | 1.000 | [0.935, 1.070] | 0.997 |  | -0.011 | [-0.043, 0.020] | 0.491 |
| SSRI non-response | 0.977 | [0.912, 1.046] | 0.501 |  | 0.030 | [-0.002, 0.062] | 0.062 |
| Treatment resistant depression | 0.980 | [0.871, 1.104] | 0.743 |  | 0.024 | [-0.031, 0.079] | 0.397 |
| Lithium response | 1.031 | [0.959, 1.110] | 0.407 |  | -0.011 | [-0.045, 0.023] | 0.537 |

### 8 Gene-set level analyses

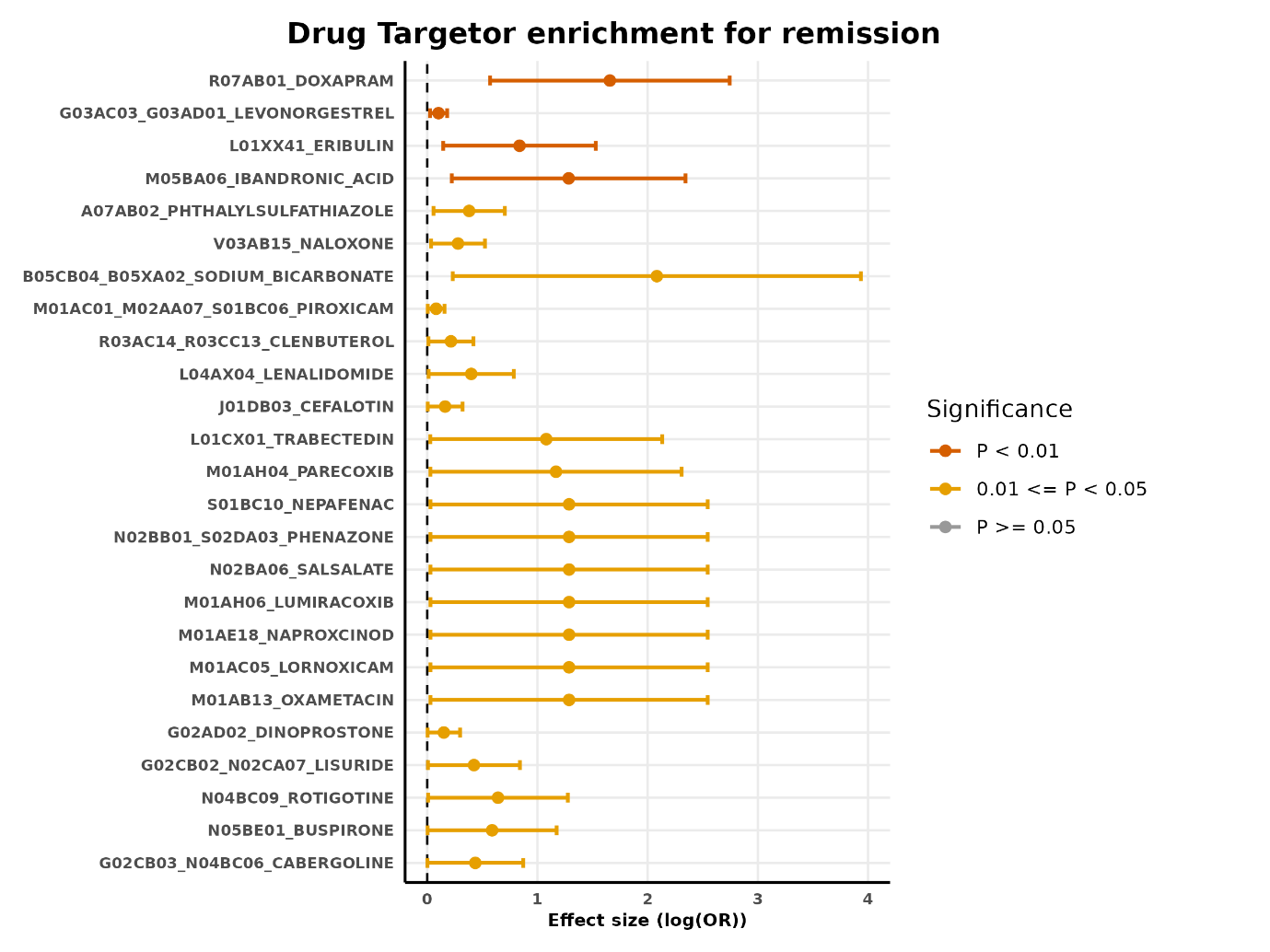

Figure S8 MAGMA drug enrichment analysis for non-remission

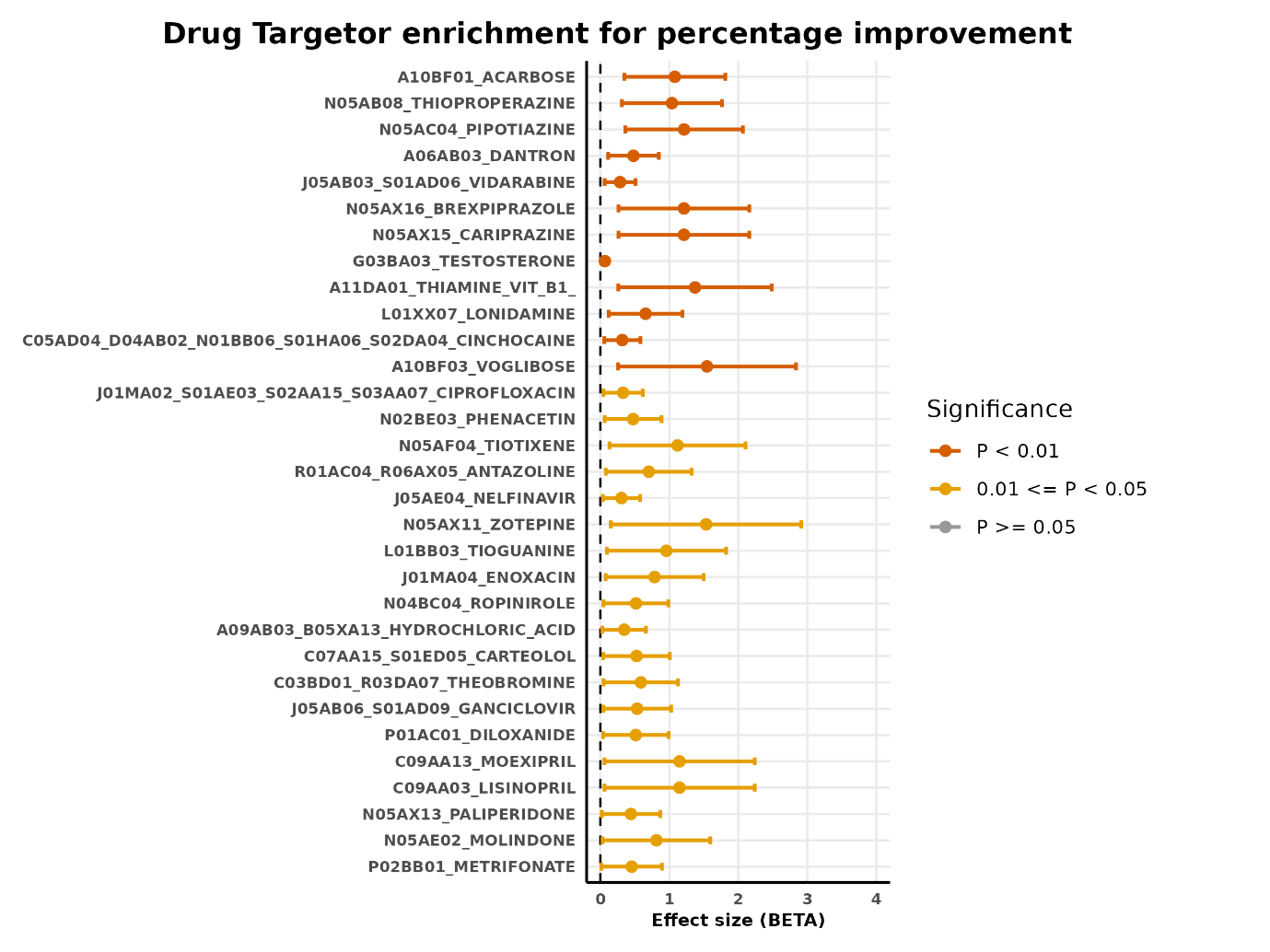

Figure S9 MAGMA drug enrichment analysis for percentage improvement

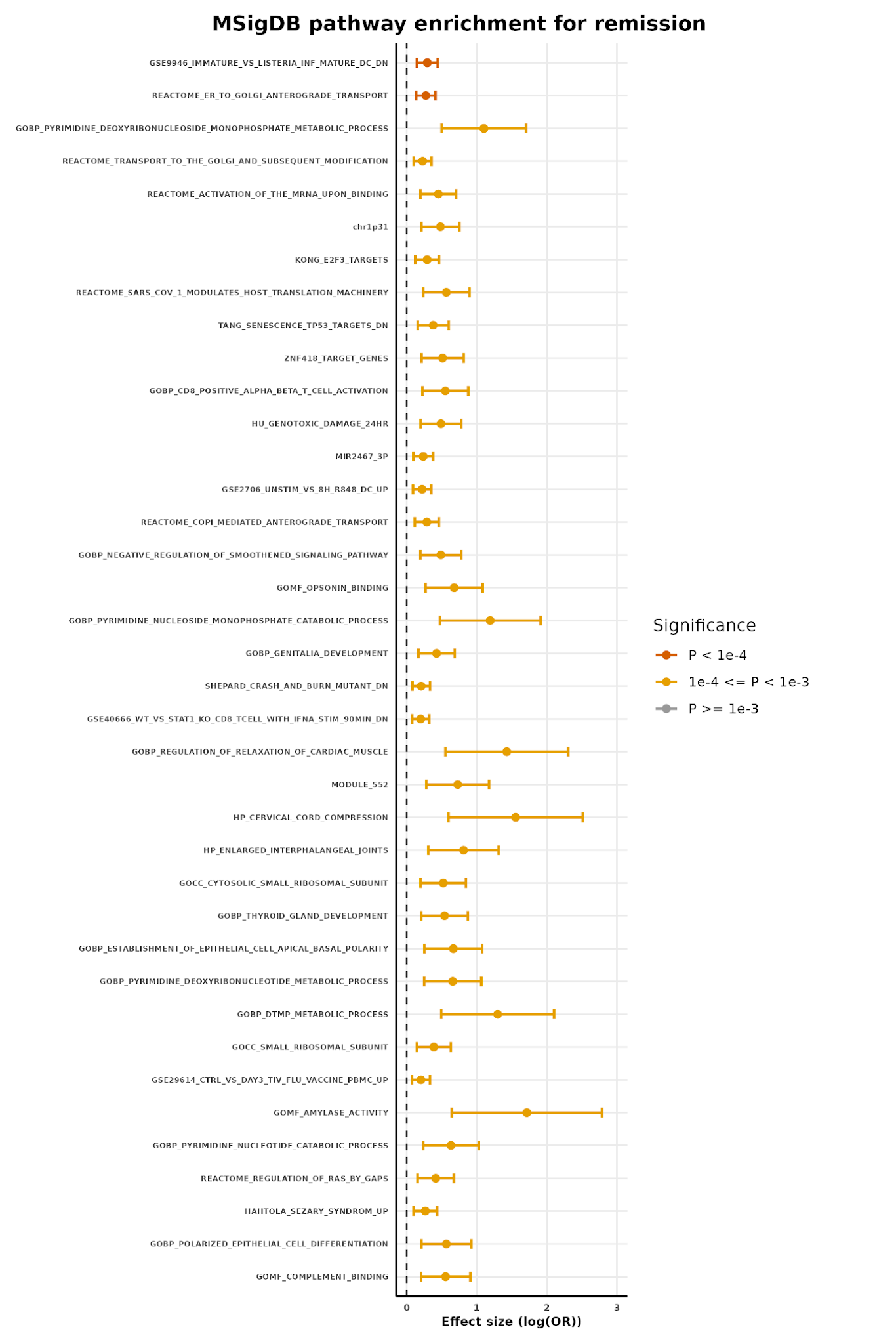

Figure S10 MAGMA MSigDB enrichment analysis for non-remission

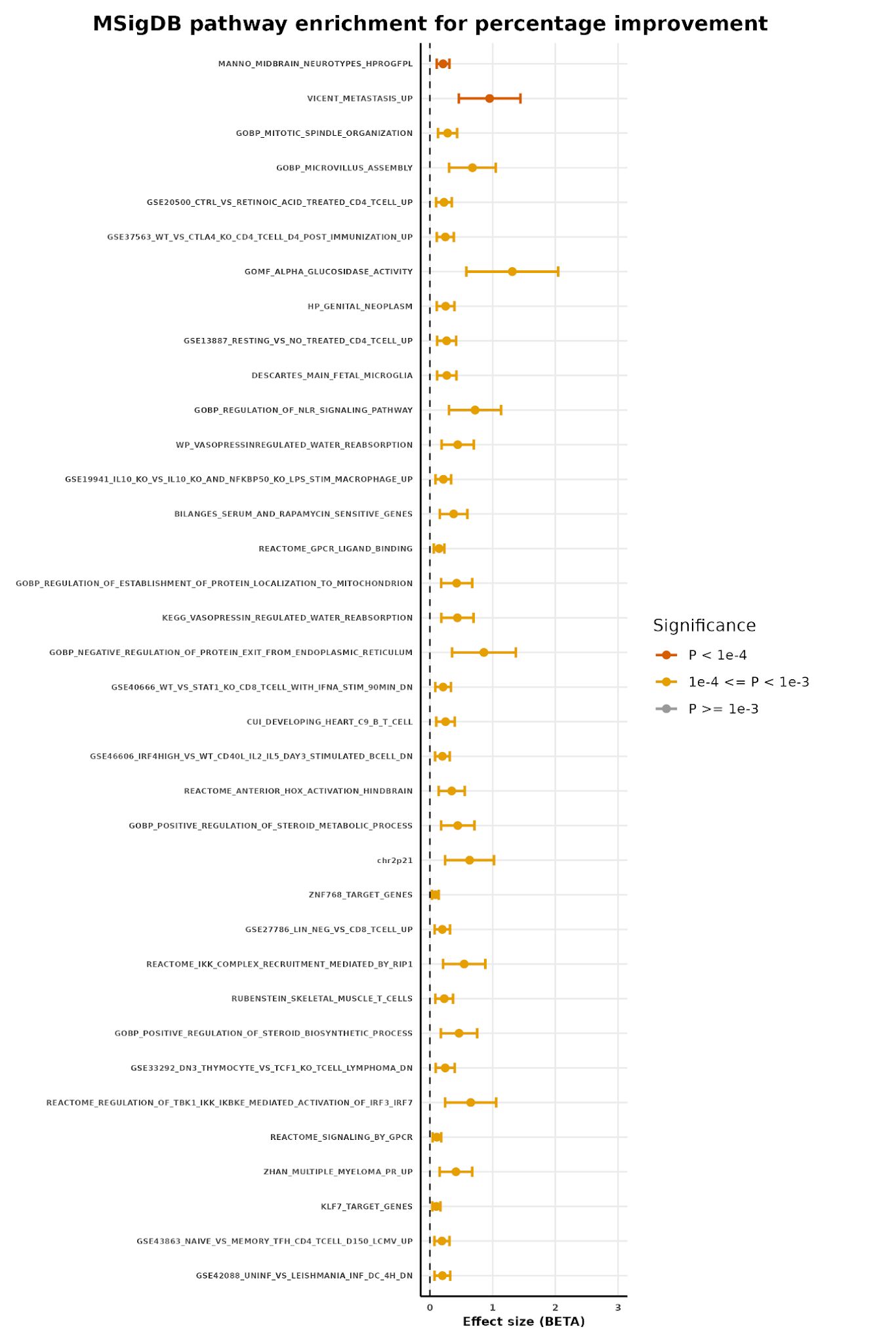

Figure S11 MAGMA MSigDB enrichment analysis for percentage improvement

**Major Depressive Disorder Working Group of the Psychiatric Genomics Consortium**

Mark J Adams* 1

Fabian Streit* 2, 3, 4, 5

Xiangrui Meng* 6

Swapnil Awasthi* 7

Brett N Adey 8

Karmel W Choi 9, 10

V Kartik Chundru 11, 12

Jonathan RI Coleman 8, 13

Bart Ferwerda 14

Jerome C Foo 2, 15, 16, 17

Zachary F Gerring 18

Olga Giannakopoulou 6

Priya Gupta 19, 20

Alisha S M Hall 2, 21

Arvid Harder 22

David M Howard 8

Christopher Hübel 8, 23, 24

Alex S F Kwong 1, 25

Daniel F Levey 19, 20

Brittany L Mitchell 18, 26, 27, 28

Guiyan Ni 29

Vanessa K Ota 30

Oliver Pain 31

Gita A Pathak 19, 32

Eva C Schulte 33, 34, 35, 36, 37

Xueyi Shen 1

Jackson G Thorp 18

Alicia Walker 29

Shuyang Yao 22

Jian Zeng 29

Johan Zvrskovec 8, 13

Dag Aarsland 38

Ky'Era V Actkins 39

Mazda Adli 40, 41

Esben Agerbo 24, 42, 43

Mareike Aichholzer 44

Allison Aiello 45

Tracy M Air 46

Thomas D Als 43, 47, 48

Evelyn Andersson 49

Till F M Andlauer 50, 51

Volker Arolt 52

Helga Ask 53, 54

Julia Bäckman 49

Sunita Badola 55

Clive Ballard 56

Karina Banasik 57

Nicholas J Bass 6

Aartjan T F Beekman 58

Sintia Belangero 30, 59

Tim B Bigdeli 60

Elisabeth B Binder 50, 61, 62

Ottar Bjerkeset 63, 64

Gyda Bjornsdottir 65

Sigrid Børte 66, 67, 68

Emma Bränn 69

Alice Braun 7

Thorsten Brodersen 70

Tanja M Brückl 71

Søren Brunak 57

Mie T Bruun 72

Margit Burmeister 73

Pichit Buspavanich 74, 75

Jonas Bybjerg-Grauholm 76, 77

Enda M Byrne 78

Jianwen Cai 79

Archie Campbell 80, 81

Megan L Campbell 82

Adrian I Campos 83

Enrique Castelao 84

Jorge Cervilla 85, 86, 87

Boris Chaumette 88

Chia-Yen Chen 89

Hsi-Chung Chen 90, 91

Zhengming Chen 92

Sven Cichon 93, 94, 95, 96

Lucía Colodro-Conde 18, 97

Anne Corbett 56

Elizabeth C Corfield 53, 98

Baptiste Couvy-Duchesne 99

Nick Craddock 100

Udo Dannlowski 52

Gail Davies 101

EJC de Geus 102

Ian J Deary 101

Franziska Degenhardt 94, 103

Abbas Dehghan 104, 105

J Raymond DePaulo 106

Michael Deuschle 5, 107

Maria Didriksen 108

Khoa Manh Dinh 109

Nese Direk 110

Srdjan Djurovic 111, 112

Anna R Docherty 113, 114, 115

Katharina Domschke 116

Joseph Dowsett 108

Ole Kristian Drange 63, 117, 118, 119

Erin C Dunn 10, 120, 121

William Eaton 122

Gudmundur Einarsson 65

Thalia C Eley 8

Samar S M Elsheikh 123

Jan Engelmann 124

Michael E Benros 77, 125, 126

Christian Erikstrup 109

Valentina Escott-Price 100

Chiara Fabbri 8, 127

Yu Fang 73

Sarah Finer 128

Josef Frank 2

Robert C Free 129

Linda Gallo 130

He Gao 131

Michael Gill 132

Maria Gilles 5, 107

Fernando S Goes 106

Scott Douglas Gordon 18, 26

Jakob Grove 43, 47, 48, 133

Daniel F Gudbjartsson 65, 134

Blanca Gutierrez 85, 86, 87

Tim Hahn 52

Lynsey S Hall 1, 100

Thomas F Hansen 57, 135, 136

Magnus Haraldsson 137, 138

Catharina A Hartman 139

Alexandra Havdahl 53, 140

Caroline Hayward 141

Stefanie Heilmann-Heimbach 94

Stefan Herms 93, 94

Ian B Hickie 142

Henrik Hjalgrim 143

Jens Hjerling-Leffler 144

Per Hoffmann 93, 94

Georg Homuth 145

Carsten Horn 146

Jouke-Jan Hottenga 102

David M Hougaard 76, 77

Iiris Hovatta 147

Qin Qin Huang 12

Donald Hucks 39

Floris Huider 102

Karen A Hunt 148

Nicholas S Ialongo 122

Marcus Ising 149

Erkki Isometsä 150

Rick Jansen 58

Yunxuan Jiang 151

Ian Jones 100

Lisa A Jones 152

Lina Jonsson 153

Masahiro Kanai 154, 155, 156

Robert Karlsson 22

Siegfried Kasper 157

Kenneth S Kendler 158

Ronald C Kessler 159

Stefan Kloiber 123, 149, 160, 161

James A Knowles 162

Nastassja Koen 82

Julia Kraft 7

Henry R Kranzler 163, 164

Kristi Krebs 165

Theodora Kunovac Kallak 166

Zoltán Kutalik 167, 168, 169

Elisa Lahtela 170

Marilyn Lake 171

Margit Hørup Larsen 108

Eric J Lenze 172

Melissa Lewins 1

Glyn Lewis 6

Liming Li 173, 174

Bochao Danae Lin 175

Kuang Lin 92

Penelope A Lind 18, 26, 27, 28

Yu-Li Liu 176

Donald J MacIntyre 1

Dean F MacKinnon 106

Brion S Maher 122

Wolfgang Maier 177

Victoria S Marshe 123, 178

Gabriela A Martinez-Levy 179

Koichi Matsuda 180, 181

Hamdi Mbarek 102

Peter McGuffin 8

Sarah E Medland 18, 26, 182, 183

Susanne Meinert 52, 184

Christina Mikkelsen 108, 185

Susan Mikkelsen 109

Yuri Milaneschi 58

Iona Y Millwood 92

Esther Molina 86, 87, 186

Francis M Mondimore 106

Preben Bo Mortensen 24, 42, 43

Benoit H Mulsant 123, 160

Joonas Naamanka 147

Jake M Najman 187

Matthias Nauck 188, 189

Igor Nenadić 190

Kasper R Nielsen 191

Ilja M Nolte 192

Merete Nordentoft 77, 125, 126

Markus M Nöthen 94

Mette Nyegaard 43, 76, 193

Michael C O'Donovan 100

Asmundur Oddsson 65

Adrielle M Oliveira 194

Catherine M Olsen 195, 196

Hogni Oskarsson 197

Sisse Rye Ostrowski 108, 198

Michael J Owen 100

Richard Packer 199

Teemu Palviainen 170

Pedro M Pan 194

Carlos N Pato 200

Michele T Pato 200

Nancy L Pedersen 22

Ole Birger Pedersen 70, 198

Wouter J Peyrot 58

James B Potash 106

Martin Preisig 84

Michael H Preuss 201, 202

Jorge A Quiroz 203

Miguel E Renteria 18, 27, 28

Charles F Reynolds III 204

John P Rice 172

Saori Sakaue 154, 155, 205

Marcos L Santoro 206

Robert A Schoevers 207, 208

Andrew Schork 43, 209

Thomas G Schulze 2, 34, 106, 210, 211, 212

Tabea S Send 107

Jianxin Shi 213

Engilbert Sigurdsson 214

Kritika Singh 39

Grant C B Sinnamon 215

Lea Sirignano 2, 5

Olav B Smeland 119, 216

Daniel J Smith 217

Tamar Sofer 218

Erik Sørensen 108

Sundararajan Srinivasan 219

Hreinn Stefansson 65

Kari Stefansson 65, 138

Peter Straub 39

Mei-Hsin Su 220

André Tadic 124, 221

Henning Teismann 222

Alexander Teumer 223

Anita Thapar 100, 224

Pippa A Thomson 81

Lise Wegner Thørner 108

Apostolia Topaloudi 225

Shih-Jen Tsai 226, 227

Ioanna Tzoulaki 104, 105, 228

George Uhl 229

André G Uitterlinden 230

Henrik Ullum 108, 198, 231

Daniel Umbricht 232

Robert J Ursano 233

Sandra Van der Auwera 223

Albert M van Hemert 234

Abirami Veluchamy 219

Alexander Viktorin 22

Henry Völzke 235

G Bragi Walters 65

Xiaotong Wang 236

Agaz Wani 237

Myrna M Weissman 238

Jürgen Wellmann 222

David C Whiteman 195

Derek Wildman 237

Gonneke Willemsen 102

Alexander T Williams 199

Bendik S Winsvold 67, 68, 239

Stephanie H Witt 2, 5, 240

Ying Xiong 22

Lea Zillich 2

John-Anker Zwart 66, 67, 68

23andMe Research Team 151

China Kadoorie Biobank Collaborative Group 241

Estonian Biobank Research Team 165

Genes & Health Research Team 242

HUNT All-In Psychiatry 243

The BioBank Japan Project 244

VA Million Veteran Program 245

Ole A Andreassen 119, 216, 246

Bernhard T Baune 247, 248, 249

Klaus Berger 222

Dorret I Boomsma 102, 250

Anders D Børglum 43, 47, 48

Gerome Breen 8, 13

Na Cai 251, 252, 253

Hilary Coon 113, 115

William E Copeland 254

Byron Creese 56

Carlos S Cruz-Fuentes 179

Darina Czamara 71

Lea K Davis 39, 255

Eske M Derks 18

Enrico Domenici 256

Paul Elliott 104, 105, 228, 257

Andreas J Forstner 94, 96, 258

Micha Gawlik 259

Joel Gelernter 19, 20, 260

Hans J Grabe 223

Steven P Hamilton 261

Kristian Hveem 67, 262, 263

Catherine John 199, 264

Jaakko Kaprio 170

Tilo Kircher 190

Marie-Odile Krebs 265

Po-Hsiu Kuo 90, 266

Mikael Landén 22, 153

Kelli Lehto 165

Douglas F Levinson 267

Qingqin S Li 268

Klaus Lieb 124

Ruth J F Loos 185, 201, 269, 270, 271

Yi Lu 22

Susanne Lucae 149

Jurjen J Luykx 58, 175, 272

Hermine HM Maes 158, 220, 273

Patrik K Magnusson 22

Hilary C Martin 12

Nicholas G Martin 18, 26

Andrew McQuillin 6

Christel M Middeldorp 78, 274

Lili Milani 165

Ole Mors 43, 275

Daniel J Müller 123, 160, 161, 276

Bertram Müller-Myhsok 50, 277, 278

Yukinori Okada 154, 279, 280

Albertine J Oldehinkel 139

Sara A Paciga 281

Colin NA Palmer 219

Peristera Paschou 225

Brenda WJH Penninx 58

Roy H Perlis 9, 10, 282

Roseann E Peterson 60

Giorgio Pistis 84

Renato Polimanti 19, 32

David J Porteous 81

Danielle Posthuma 283, 284

Jill A Rabinowitz 285

Ted Reichborn-Kjennerud 53

Andreas Reif 44

Frances Rice 100, 224

Roland Ricken 7

Marcella Rietschel 2

Margarita Rivera 86, 87, 286

Christian Rück 49

Giovanni A Salum 287

Catherine Schaefer 288

Srijan Sen 73, 289

Alessandro Serretti 290, 291

Alkistis Skalkidou 166

Jordan W Smoller 9, 292, 293

Dan J Stein 82

Frederike Stein 294

Murray B Stein 295, 296, 297, 298

Patrick F Sullivan 22, 299

Martin Tesli 300

Thorgeir E Thorgeirsson 65

Henning Tiemeier 301, 302

Nicholas J Timpson 25, 303

Monica Uddin 237

Rudolf Uher 304

David A van Heel 148

Karin JH Verweij 305

Robin G Walters 92

Sylvia Wassertheil-Smoller 306

Jens R Wendland 55

Thomas Werge 77, 198, 209, 307, 308

Aeilko H Zwinderman 14

Karoline Kuchenbaecker* 6, 92

Naomi R Wray* 29, 236, 309

Stephan Ripke* 7, 293

Cathryn M Lewis* 8, 310

Andrew M McIntosh* 1, 81

***** shared first and last authors

V. 2024-09-24 22:50

1, Division of Psychiatry, University of Edinburgh, Edinburgh, UK

2, Department of Genetic Epidemiology in Psychiatry, Central Institute of Mental Health, Medical Faculty Mannheim, Heidelberg University, Mannheim, BW, DE

3, Hector Institute for Artificial Intelligence in Psychiatry, Central Institute of Mental Health, Medical Faculty Mannheim, Heidelberg University, Mannheim, BW, DE

4, Department for Psychiatry and Psychotherapy, Central Institute of Mental Health, Medical Faculty Mannheim, Heidelberg University, Mannheim, BW, DE

5, German Center for Mental Health (DZPG), Partner Site Mannheim - Heidelberg - Ulm, DE

6, Division of Psychiatry, University College London, London, UK

7, Department of Psychiatry and Psychotherapy, Charité – Universitätsmedizin Berlin, Berlin, BE, DE

8, Social, Genetic and Developmental Psychiatry Centre, King's College London, London, UK

9, Department of Psychiatry, Massachusetts General Hospital, Boston, MA, US

10, Department of Psychiatry, Harvard Medical School, Boston, MA, US

11, Department of Clinical and Biomedical Sciences, Faculty of Health and Life Sciences, University of Exeter, Exeter, UK

12, Human Genetics, Wellcome Sanger Institute, Hinxton, UK

13, NIHR Maudsley Biomedical Research Centre, King's College London, London, UK

14, Epidemiologie en Data Science (EDS), Amsterdam UMC, location University of Amsterdam, Amsterdam, NL

15, Institute for Psychopharmacology, Central Institute of Mental Health, Medical Faculty Mannheim, Heidelberg University, Mannheim, BW, DE

16, Department of Psychiatry, College of Health Sciences, University of Alberta, Edmonton, AB, CA

17, Neuroscience and Mental Health Institute, University of Alberta, Edmonton, AB, CA

18, Brain & Mental Health Program, QIMR Berghofer Medical Research Institute, Brisbane, QLD, AU

19, Department of Psychiatry, Yale University School of Medicine, New Haven, CT, US

20, Department of Psychiatry, Veterans Affairs Connecticut Healthcare System, West Haven, CT, US

21, Department of Clinical Medicine, Aarhus University, Aarhus, DK

22, Department of Medical Epidemiology and Biostatistics, Karolinska Institutet, Stockholm, SE

23, Department of Pediatric Neurology, Charité – Universitätsmedizin Berlin, Berlin, BE, DE

24, National Centre for Register-based Research, Aarhus University, Aarhus, DK

25, MRC Integrative Epidemiology Unit, University of Bristol, Bristol, UK

26, Mental Health and Neuroscience, QIMR Berghofer Medical Research Institute, Brisbane, QLD, AU

27, School of Biomedical Sciences, Queensland University of Technology, Brisbane, QLD, AU

28, School of Biomedical Sciences, The University of Queensland, Brisbane, QLD, AU

29, Institute for Molecular Bioscience, University of Queensland, Brisbane, QLD, AU

30, Morphology and Genetics, Universidade Federal de Sao Paulo, Sao Paulo, SP, BR

31, Maurice Wohl Clinical Neuroscience Institute, Department of Basic and Clinical Neuroscience, King's College London, London, UK

32, Veterans Affairs Connecticut Healthcare System, West Haven, CT, US

33, Department of Psychiatry and Psychotherapy, University Hospital, LMU Munich, Munich, BY, DE

34, Institute of Psychiatric Phenomics and Genomics, University Hospital, LMU Munich, Munich, BY, DE

35, Department of Psychiatry and Psychotherapy, University Hospital Bonn, Medical Faculty, University of Bonn, Bonn, DE

36, Institute of Human Genetics, University Hospital Bonn, Medical Faculty, University of Bonn, Bonn, DE

37, German Center for Mental Health (DZPG), Partner Site Munich - Augsburg, DE

38, Old Age Psychiatry, King's College London, London, UK

39, Department of Medicine, Division of Genetic Medicine, Vanderbilt University Medical Center, Nashville, TN, US

40, Department of Psychiatry and Psychotherapy, Charité – Universitätsmedizin Berlin, Campus Charité Mitte (CCM), Berlin, BE, DE

41, Center for Psychiatry, Psychotherapy and Psychosomatic Medicine, Fliedner Klinik Berlin, Berlin, BE, DE

42, Centre for Integrated Register-based Research, Aarhus University, Aarhus, DK

43, iPSYCH, The Lundbeck Foundation Initiative for Integrative Psychiatric Research, Aarhus, DK

44, Department of Psychiatry, Psychosomatic Medicine and Psychotherapy, Goethe University Frankfurt - University Hospital, Frankfurt am Main, DE

45, Department of Epidemiology, Columbia University Mailman School of Public Health, New York, NY, US

46, Discipline of Psychiatry, University of Adelaide, Adelaide, SA, AU

47, Department of Biomedicine and Centre for Integrative Sequencing, iSEQ, Aarhus University, Aarhus, DK

48, Center for Genomics and Personalized Medicine, Aarhus University, Aarhus, DK

49, Department of Clinical Neuroscience, Karolinska Institutet, Stockholm, SE

50, Department of Translational Research in Psychiatry, Max Planck Institute of Psychiatry, Munich, BY, DE

51, Department of Neurology, Klinikum rechts der Isar, Technical University of Munich, Munich, BY, DE

52, Institute for Translational Psychiatry, University of Münster, Münster, NRW, DE

53, PsychGen Centre for Genetic Epidemiology and Mental Health, Norwegian Institute of Public Health, Oslo, OSL, NO

54, PROMENTA Research Center, Department of Psychology, University of Oslo, Oslo, OSL, NO

55, Research and Development, Takeda Pharmaceutical Company Limited, Cambridge, MA, US

56, Faculty of Health and Life Sciences, University of Exeter, Exeter, UK

57, Novo Nordisk Foundation Center for Protein Research, Faculty of Health and Medical Sciences, University of Copenhagen, Copenhagen, CPH, DK

58, Department of Psychiatry, Amsterdam Public Health and Amsterdam Neuroscience, Amsterdam UMC, Vrije Universiteit Amsterdam, Amsterdam, NL

59, Laboratory of Integrative Neuroscience, Universidade Federal de Sao Paulo, Sao Paulo, SP, BR

60, Department of Psychiatry and Behavioral Sciences, Institute for Genomics in Health, State University of New York Downstate Health Sciences University, Brooklyn, NY, US

61, Department of Psychiatry and Behavioral Sciences, Emory University School of Medicine, Atlanta, GA, US

62, Department Genes and Environment, Max Planck Institute of Psychiatry, Munich, BY, DE

63, Department of Mental Health, Faculty of Medicine and Health Sciences, Norwegian University of Science and Technology (NTNU), Trondheim, TRD, NO

64, Faculty of Nursing and Health Sciences, NORD University, Levanger, NO

65, deCODE Genetics / Amgen, Reykjavik, IS

66, Institute of Clinical Medicine, Faculty of Medicine, University of Oslo, Oslo, OSL, NO

67, HUNT Center for Molecular and Clinical Epidemiology, Department of Public Health and Nursing, Faculty of Medicine and Health Sciences, Norwegian University of Science and Technology, Trondheim, TRD, NO

68, Department of Research and Innovation, Division of Clinical Neuroscience, Oslo University Hospital, Oslo, OSL, NO

69, Institute of Environmental Medicine, Unit of Integrative Epidemiology, Karolinska Institutet, Stockholm, SE

70, Department of Clinical Immunology, Zealand University Hospital, Køge, DK

71, Department Genes and Environment, Max Planck Institute of Psychiatry, Munich, BY, DE

72, Department of Clinical Immunology, Odense University Hospital, Odense, DK

73, Michigan Neuroscience Institute, University of Michigan, Ann Arbor, MI, US

74, Department of Psychiatry and Psychotherapy, Gender Research in Medicine, Institute of Sexology and Sexual Medicine, Charité – Universitätsmedizin Berlin, Berlin, BE, DE

75, Department of Psychiatry, Psychotherapy and Psychosomatics, Brandenburg Medical School Theodor Fontane, Neuruppin, BB, DE

76, Center for Neonatal Screening, Department for Congenital Disorders, Statens Serum Institut, Copenhagen, CPH, DK

77, iPSYCH, The Lundbeck Foundation Initiative for Integrative Psychiatric Research, Copenhagen, CPH, DK

78, Child Health Research Centre, University of Queensland, Brisbane, QLD, AU

79, Department of Biostatistics , University of North Carolina at Chapel Hill  , Chapel Hill, NC, US

80, Centre for Medical Informatics, Usher Institute, University of Edinburgh, Edinburgh, UK

81, Centre for Genomic & Experimental Medicine, Institute for Genetics and Cancer, University of Edinburgh, Edinburgh, UK

82, SAMRC Unit on Risk & Resilience in Mental Disorders, Department of Psychiatry and Neuroscience Institute, University of Cape Town, Cape Town, SA

83, Statistical genetics, Institute for Molecular Bioscience, The University of Queensland, Brisbane, QLD, AU

84, Department of Psychiatry, Lausanne University Hospital and University of Lausanne, Prilly, VD, CH

85, Department of Psychiatry, Faculty of Medicine, University of Granada, Granada, ES

86, Instituto de Investigación Biosanitaria, Ibs Granada, Granada, ES

87, Institute of Neurosciences ´Federico Olóriz´, Biomedical Research Centre (CIBM), University of Granada, Granada, ES

88, Université de Paris Cité, INSERM U1266, Institute of Psychiatry and Neuroscience of Paris, GHU Paris Psychiatry and Neuroscience, Paris, FR

89, Biogen, Cambridge, MA, US

90, Department of Psychiatry, National Taiwan University Hospital,, TW

91, School of Medicine, National Taiwan University College of Medicine, Taipei, TW

92, Nuffield Department of Population Health, University of Oxford, Oxford, UK

93, Human Genomics Research Group, Department of Biomedicine, University of Basel, Basel, CH

94, Institute of Human Genetics, University of Bonn, School of Medicine & University Hospital Bonn, Bonn, DE

95, Institute of Medical Genetics and Pathology, University Hospital Basel, University of Basel, Basel, CH

96, Institute of Neuroscience and Medicine (INM-1), Research Center Juelich, Juelich, DE

97, School of Psychology, University of Queensland, Brisbane, QLD, AU

98, Nic Waals Institute, Lovisenberg Diakonale Hospital, Oslo, OSL, NO

99, Centre for Advanced Imaging, University of Queensland, Saint Lucia, QLD, AU

100, Centre for Neuropsychiatric Genetics and Genomics, Cardiff University, Cardiff, UK

101, The Lothian Birth Cohorts, University of Edinburgh, Edinburgh, UK

102, Department of Biological Psychology & Amsterdam Public Health Research Institute, Vrije Universiteit Amsterdam, Amsterdam, NL

103, Department of Child and Adolescent Psychiatry, Psychosomatics and Psychotherapy, University Hospital Essen, University of Duisburg-Essen, Duisburg, DE

104, MRC Centre for Environment and Health, School of Public Health, Imperial College London, London, UK

105, Imperial College Dementia Research Institute, Imperial College London, London, UK

106, Department of Psychiatry and Behavioral Sciences, Johns Hopkins University School of Medicine, Baltimore, MD, US

107, Department of Psychiatry and Psychotherapy, Research Group Stress Related Disorders, Central Institute of Mental Health, Medical Faculty Mannheim, Heidelberg University, Mannheim, BW, DE

108, Department of Clinical Immunology, Copenhagen University Hospital, Rigshospitalet, Copenhagen, CPH, DK

109, Department of Clinical Immunology, Aarhus University Hospital, Aarhus, DK

110, Department of Psychiatry, Istanbul University, Istanbul, TR

111, Department of Medical Genetics, Oslo University Hospital, Oslo, OSL, NO

112, NORMENT, Department of Clinical Science, University of Bergen, Bergen, NO

113, Psychiatry, University of Utah School of Medicine, Salt Lake City, UT, US

114, Center for Genomic Medicine, Salt Lake City, UT, US

115, Huntsman Mental Health Institute, Salt Lake City, UT, US

116, Department of Psychiatry and Psychotherapy, Medical Center, University of Freiburg, Faculty of Medicine, University of Freiburg, Freiburg, DE

117, Division of Mental Health Care, St. Olavs Hospital, Trondheim University Hospital, Trondheim, TRD, NO

118, Department of Psychiatry, Sørlandet Hospital, Kristiansand, AG, NO

119, NORMENT, Institute of Clinical Medicine, University of Oslo, Oslo, OSL, NO

120, Center for Genomic Medicine, Massachusetts General Hospital, Boston, MA, US

121, Department of Sociology, Purdue University, West Lafayette, IN, US

122, Department of Mental Health, Johns Hopkins, Baltimore, MD, US

123, Centre for Addiction and Mental Health, Toronto, ON, CA

124, Department of Psychiatry and Psychotherapy, University Medical Center of the Johannes Gutenberg University Mainz, Mainz, DE

125, Mental Health Center Copenhagen, Mental Health Services Capital Region of Denmark, Copenhagen, CPH, DK

126, Faculty of Health Science, Department of Clinical Medicine, University of Copenhagen, Copenhagen, CPH, DK

127, Department of Biomedical and Neuromotor Sciences, University of Bologna, Bologna, IT

128, Wolfson Institute of Population Health, Queen Mary University of London, London, UK

129, School of Computing and Mathematical Sciences, University of Leicester, Leicester, UK

130, Department of Psychology, San Diego San Diego State University, San Diego, CA, US

131, Department of Epidemiology and Biostatistics, Imperial College London, London, UK

132, Discipline of Psychiatry, School of Medicine, Trinity College Dublin, Dublin, IE

133, Bioinformatics Research Centre, Aarhus University, Aarhus, DK

134, School of Engineering, University of Iceland, Reykjavik, IS

135, Danish Headache Centre, Department of Neurology, Rigshospitalet, Glostrup, DK

136, Neurogenomics Group, Translational Research Centre, Rigshospitalet Copenhagen University Hospital, Glostrup, DK

137, Landspitali-University Hospital, Reykjavik, IS

138, Faculty of Medicine, University of Iceland, Reykjavik, IS

139, Department of Psychiatry, University of Groningen, University Medical Center Groningen, Groningen, NL

140, Nic Waals Institute, Lovisenberg Diaconal Hospital, Oslo, OSL, NO

141, MRC Human Genetics Unit, Institute for Genetics and Cancer, University of Edinburgh, Edinburgh, UK

142, Brain and Mind Centre, University of Sydney, Sydney, NSW, AU

143, Department of Epidemiology Research, Statens Serum Institut, Copenhagen, CPH, DK

144, Department of Medical Biochemistry and Biophysics, Karolinska Institutet, Stockholm, SE

145, Interfaculty Institute for Genetics and Functional Genomics, Department of Functional Genomics, University Medicine Greifswald, Greifswald, MV, DE

146, Roche Pharmaceutical Research and Early Development, Pharmaceutical Sciences, Roche Innovation Center Basel, F. Hoffmann-La Roche Ltd, Basel, CH

147, SleepWell Research Program and Department of Psychology and Logopedics, University of Helsinki, Helsinki, FI

148, Blizard Institute, Barts and the London School of Medicine and Dentistry, Queen Mary University of London, London, UK

149, Max Planck Institute of Psychiatry, Munich, BY, DE

150, Department of Psychiatry, University of Helsinki, Helsinki, FI

151, 23andMe Research Team, 23andMe, Inc., Sunnyvale, CA, US

152, Department of Psychological Medicine, University of Worcester, Worcester, UK

153, Institution of Neuroscience and Physiology, University of Gothenburg, Gothenburg, SE

154, Department of Statistical Genetics, Osaka University Graduate School of Medicine, Suita, JP

155, Program in Medical and Population Genetics, Broad Institute of Harvard and MIT, Cambridge, MA, US

156, Center for Computational and Integrative Biology, Massachusetts General Hospital, Boston, MA, US

157, Center for Brain Research, Department of Molecular Neuroscience, Medical University of Vienna, Vienna, AT

158, Department of Psychiatry, Virginia Commonwealth University, Richmond, VA, US

159, Department of Health Care Policy, Harvard Medical School, Boston, MA, US

160, Department of Psychiatry, University of Toronto, Toronto, ON, CA

161, Department of Pharmacology & Toxicology, University of Toronto, Toronto, ON, CA

162, Department of Genetics, Rutgers University, Piscataway, NJ, US

163, Department of Psychiatry, Perelman School of Medicine, University of Pennsylvania, Philadelphia, PA, US

164, Mental Illness Research, Education and Clinical Center, Crescenz VA Medical Center, Philadelphia, PA, US

165, Estonian Genome Centre, Institute of Genomics, University of Tartu, Tartu, EE

166, Department of Women's and Children's Health, Uppsala University, Uppsala, SE

167, Department of Epidemiology and Health Systems, Center for Primary Care and Public Health, Lausanne, VD, CH

168, Department of Computational Biology, University of Lausanne, Lausanne, VD, CH

169, Swiss Institute of Bioinformatics, Lausanne, VD, CH

170, Institute for Molecular Medicine Finland - FIMM, University of Helsinki, Helsinki, FI

171, SAMRC Unit on Risk & Resilience in Mental Disorders, Department of Psychiatry and Neuroscience Institute, University of Cape Town, Cape Town, SA

172, Department of Psychiatry, Washington University School of Medicine in St. Louis, St. Louis, MO, US

173, Department of Epidemiology and Biostatistics, School of Public Health, Peking University, Beijing, CN

174, Peking University Center for Public Health and Epidemic Preparedness & Response, Peking University, Beijing, CN

175, Department of Psychiatry and Neuropsychology, School for Mental Health and Neuroscience, Maastricht University Medical Centre, Maastricht, NL

176, Center for Neuropsychiatric Research, National Health Research Institutes,, TW

177, Department of Psychiatry and Psychotherapy, University of Bonn, Bonn, DE

178, Center for Translational and Computational Neuroimmunology, Columbia University Medical Center, New York, NY, US

179, Psychiatric Genetics Department, Instituto Nacional de Psiquiatría Ramón de la Fuente Muñiz (INPRFM), Mexico City, CDMX, MX

180, Laboratory of Genome Technology, Human Genome Center, Institute of Medical Science, The University of Tokyo, Tokyo, JP

181, Laboratory of Clinical Genome Sequencing, Department of Computational Biology and Medical Sciences, Graduate School of Frontier Sciences, The University of Tokyo, Tokyo, JP

182, School of Psychology, The University of Queensland, Brisbane, QLD, AU

183, School of Psychology and Counselling, Queensland University of Technology, Brisbane, QLD, AU

184, Institute for Translational Neuroscience, University of Münster, Münster, NRW, DE

185, Novo Nordisk Foundation Center for Basic Metabolic Research, Faculty of Health and Medical Sciences, University of Copenhagen, Copenhagen, CPH, DK

186, Department of Nursing, Faculty of Health Sciences, University of Granada, Granada, ES

187, School of Public Health, University of Queensland, Brisbane, QLD, AU

188, Institute of Clinical Chemistry and Laboratory Medicine, University Medicine Greifswald, Greifswald, MV, DE

189, DZHK (German Centre for Cardiovascular Research), Partner Site Greifswald, Greifswald, MV, DE

190, Department of Psychiatry, University of Marburg, Marburg, HE, DE

191, Department of Clinical Immunology, Aalborg University Hospital, Aalborg, DK

192, Department of Epidemiology, University of Groningen, University Medical Center Groningen, Groningen, NL

193, Department of Health, Science and Technology, Aalborg University, Aalborg, DK

194, Department of Psychiatry, Universidade Federal de Sao Paulo, Sao Paulo, SP, BR

195, Population Health, QIMR Berghofer Medical Research Institute, Brisbane, QLD, AU

196, The Fraser Institute, Faculty of Medicine, University of Queensland, Brisbane, QLD, AU

197, Directorate of Health, Iceland, Reykjavik, IS

198, Department of Clinical Medicine, University of Copenhagen, Copenhagen, CPH, DK

199, Department of Population Health Sciences, University of Leicester, Leicester, UK

200, Department of Psychiatry, Rutgers University, Piscataway, NJ, US

201, Charles Bronfman Institute for Personalized Medicine, Icahn School of Medicine at Mount Sinai, New York, NY, US

202, Department of Environmental Medicine and Public Health, Icahn School of Medicine at Mount Sinai, New York, NY, US

203, Translational Medicine, Roche, New York, NY, US

204, Psychiatry, University of Pittsburgh Medical Centre, Pittsburgh, PA, US

205, Divisions of Genetics and Rheumatology, Department of Medicine, Brigham and Women’s Hospital, Harvard Medical School, Boston, MA, US

206, Department of Biochemistry, Universidade Federal de Sao Paulo, Sao Paulo, SP, BR

207, Department of Psychiatry, University Medical Center Groningen, Groningen, NL

208, Research School of Behavioural and Cognitive Neurosciences (BCN), University of Groningen, Groningen, NL

209, Institute of Biological Psychiatry, Mental Health Center Sct. Hans, Mental Health Services Capital Region of Denmark, Copenhagen, CPH, DK

210, Department of Psychiatry and Psychotherapy, University Medical Center Göttingen, Goettingen, NI, DE

211, Human Genetics Branch, NIMH Division of Intramural Research Programs, Bethesda, MD, US

212, Department of Psychiatry and Behavioral Sciences, SUNY Upstate Medical University, Syracuse, NY, USA

213, Division of Cancer Epidemiology and Genetics, National Cancer Institute, Bethesda, MD, US

214, Faculty of Medicine, Department of Psychiatry, University of Iceland, Reykjavik, IS

215, School of Medicine and Dentistry, James Cook University, Townsville, QLD, AU

216, Division of Mental Health and Addiction, Oslo University Hospital, Oslo, OSL, NO

217, Center for Clinical Brain Sciences, University of Edinburgh, Edinburgh, UK

218, Beth Israel Deaconess Medica, Harvard Medical School, Boston, MA, US

219, Division of Population Health and Genomics, Ninewells Hospital and School of Medicine, University of Dundee, Dundee, UK

220, Virginia Institute for Psychiatric and Behavioral Genetics, Virginia Commonwealth University, Richmond, VA, US

221, Department of Psychiatry, Psychotherapy and Psychosomatics, Dr. Fontheim Mentale Gesundheit, Liebenburg, DE

222, Institute of Epidemiology and Social Medicine, University of Münster, Münster, NRW, DE

223, Department of Psychiatry and Psychotherapy, University Medicine Greifswald, Greifswald, MV, DE

224, Wolfson Centre for Young People's Mental Health, Division of Psychological Medicine and Clinical Neurosciences, Cardiff University, Cardiff, UK

225, Department of Biological Sciences, Purdue University, West Lafayette, IN, US

226, Institute of Brain Science & Division of Psychiatry, National Yang-Ming University,, TW

227, Department of Psychiatry, Taipei Veterans General Hospital, Taipei, ROC, TW

228, Imperial College BHF Centre for Research Excellence, Imperial College London, London, UK

229, University of Maryland School of Medicine and VA Maryland Healthcare System, Baltimore, MD, US

230, Department of Internal Medicine, Erasmus University Medical Center Rotterdam, Rotterdam, NL

231, Management Section, Statens Serum Institut, Copenhagen, CPH, DK

232, Xperimed LLC, Basel, CH

233, Department of Psychiatry, Uniformed Services University of the Health Sciences, Bethesda, MD, US

234, Department of Psychiatry, Leiden University Medical Center, Leiden, NL

235, Institute for Community Medicine, University Medicine Greifswald, Greifswald, MV, DE

236, Department of Psychiatry, University of Oxford, Oxford, UK

237, Genomics Program, University of South Florida College of Public Health, Tampa, FL, US

238, Columbia University Vagelos College of Physicians and Surgeons, New York, NY, US

239, Department of Neurology, Oslo University Hospital, Oslo, OSL, NO

240, Center for Innovative Psychiatric and Psychotherapeutic Research, Central Institute of Mental Health, Medical Faculty Mannheim, Heidelberg University, Mannheim, BW, DE

241, China Kadoorie Biobank Collaborative Group

242, Genes & Health Research Team

243, HUNT All-In Psychiatry

244, The BioBank Japan Project

245, VA Million Veteran Program

246, KG Jebsen Centre for Neurodevelopmental Research, University of Oslo, Oslo, OSL, NO

247, Department of Psychiatry, University of Münster, Münster, NRW, DE

248, Department of Psychiatry, University of Melbourne, Melbourne, VIC, AU

249, Florey Institute of Neuroscience and Mental Health, University of Melbourne, Melbourne, VIC, AU

250, Department of Complex Trait Genetics, CNCR, Vrije Universiteit Amsterdam, Amsterdam, NL

251, Helmholtz Pioneer Campus, Helmholtz Zentrum München, Neuherberg, DE

252, Computational Health Centre, Helmholtz Zentrum München, Neuherberg, DE

253, School of Medicine, Technical University of Munich, Munich, BY, DE

254, Department of Psychiatry, University of Vermont, Burlington, VT, US

255, Department of Medicine, Icahn School of Medicine at Mount Sinai, New York, NY, US

256, Department of Cellular, Computational and Integrative Biology, Università degli Studi di Trento, Trento, IT

257, Imperial College Biomedical Research Centre, Imperial College London, London, UK

258, Center for Human Genetics, University of Marburg, Marburg, HE, DE

259, Department of Psychiatry, Psychosomatics and Psychotherapy, Julius-Maximilians-Universität Würzburg, Würzburg, DE

260, Department of Genetics, Department of Neuroscience, Yale University School of Medicine, New Haven, CT, US

261, Psychiatry, Kaiser Permanente Northern California, San Francisco, CA, US

262, HUNT Research Center, Department of Public Health and Nursing, Faculty of Medicine and Health Sciences, Norwegian University of Science and Technology (NTNU), Trondheim, NO

263, Department of Research, Innovation and Education, St. Olavs Hospital, Trondheim University Hospital, Trondheim, TRD, NO

264, University Hospitals of Leicester NHS Trust, Leicester, UK

265, Pathophysiology of Psychiatric Diseases, INSERM, Univ Paris Cité, GHU Paris, Paris, FR

266, Institute of Epidemiology and Preventive Medicine & Department of Public Health, National Taiwan University,, TW

267, Department of Psychiatry & Behavioral Sciences, Stanford University, Stanford, CA, US

268, Neuroscience Therapeutic Area, Janssen Research and Development, LLC, Titusville, NJ, US

269, Mindich Child Health and Development Institute, Icahn School of Medicine at Mount Sinai, New York, NY, US

270, Department of Environmental Medicine and Public Health, Icahn School of Medicine at Mount Sinai, New York, NY, US

271, MRC Metabolic Diseases Unit, University of Cambridge Metabolic Research Laboratories, Wellcome-MRC Institute of Metabolic Science, Addenbrooke's Hospital, Cambridge, UK

272, Bipolar Disorders Outpatient Clinic, GGZ InGeest, Amsterdam, NL

273, Department of Human and Molecular Genetics, Virginia Commonwealth University, Richmond, VA, US

274, Child and Youth Mental Health Service, Children's Health Queensland Hospital and Health Service, Brisbane, QLD, AU

275, Psychosis Research Unit, Aarhus University Hospital-Psychiatry, Aarhus, DK

276, Department of Psychiatry, Psychosomatics and Psychotherapy, University Hospital of Würzburg, Würzburg, DE

277, Munich Cluster for Systems Neurology (SyNergy), Munich, BY, DE

278, University of Liverpool, Liverpool, UK

279, Department of Genome Informatics, Graduate School of Medicine, The University of Tokyo, Tokyo, JP

280, Laboratory for Systems Genetics, RIKEN Center for Integrative Medical Sciences, Yokohama, JP

281, Human Genetics and Computational Biomedicine, Pfizer Global Research and Development, Groton, CT, US

282, Centre for Quantitative Health, Massachusetts General Hospital, Boston, MA, US

283, Child and Adolescent Psychiatry, Amsterdam UMC, Vrije Universiteit Amsterdam, Amsterdam, NL

284, Complex Trait Genetics, Vrije Universiteit Amsterdam, Amsterdam, NL

285, Department of Mental Health, Johns Hopkins University, Baltimore, MD, US

286, Department of Biochemistry and Molecular Biology II, Faculty of Pharmacy, University of Granada, Granada, ES

287, Psychiatry, Universidade Federal do Rio Grande do Sul, Porto Alegre, BR

288, Division of Research, Kaiser Permanente Northern California, Oakland, CA, US

289, Eisenberg Family Depression Center, University of Michigan, Ann Arbor, MI, US

290, Department of Medicine and Surgery, Kore University of Enna, Enna, IT

291, Psychiatry, Oasi Research Institute-IRCCS, Troina, IT

292, Psychiatric and Neurodevelopmental Genetics Unit, Massachusetts General Hospital, Boston, MA, US

293, Stanley Center for Psychiatric Research, Broad Institute of MIT and Harvard, Cambridge, MA, US

294, Department of Psychiatry and Psychotherapy, University of Marburg, Marburg, HE, DE

295, Psychiatry Service, Veterans Affairs San Diego Healthcare System, San Diego, CA, US

296, School of Public Health, University of California, San Diego, La Jolla, CA, US

297, Department of Psychiatry, University of California, San Diego, La Jolla, CA, US

298, Psychiatry, Veterans Affairs San Diego Healthcare System, San Diego, CA, US

299, Departments of Genetics and Psychiatry, University of North Carolina at Chapel Hill, Chapel Hill, NC, US

300, Department of Mental Health and Suicide, Norwegian Institute of Public Health, Oslo, OSL, NO

301, Child and Adolescent Psychiatry, Erasmus University Medical Center Rotterdam, Rotterdam, NL

302, Social and Behavioral Science, Harvard T.H. Chan School of Public Health, Boston, MA, US

303, Population Health Sciences, Bristol Medical School, University of Bristol, Bristol, UK

304, Psychiatry, Dalhousie University, Halifax, NS, CA

305, Psychiatry, Amsterdam UMC, location University of Amsterdam, Amsterdam, NL

306, Epidemiology and Population Health, Albert Einstein College of Medicine, Bronx, NY, US

307, Institute of Biological Psychiatry, Mental Health Center Sct. Hans, Copenhagen University Hospital, Mental Health Services, Copenhagen, CPH, DK

308, GLOBE Institute, Lundbeck Foundation Centre for Geogenetics, University of Copenhagen, Copenhagen, CPH, DK

309, Queensland Brain Institute, University of Queensland, Brisbane, QLD, AU

310, Department of Medical & Molecular Genetics, King's College London, London, UK
